# Loneliness as a Pathway Linking Hearing Decline to Cognitive Aging: Longitudinal and Genetic Evidence

**DOI:** 10.64898/2026.04.16.26351059

**Authors:** Shuai Yang, Matthew D. Grilli, Robyn E. Wootton, Margot P. van de Weijer, Jorien L. Treur, Yann C. Klimentidis, David A. Sbarra

**Author notes:** **Correspondence should be directed to:** David A. Sbarra, PhD (Co-Corresponding Author); Department of Psychology, University of Arizona, Tucson, AZ, USA; Shuai Yang, MS (Co-Corresponding Author); Department of Epidemiology and Biostatistics, Mel & Enid Zuckerman College of Public Health, University of Arizona, Tucson, AZ, USA. **Author Contributions:** Shuai Yang had full access to all the study data and takes full responsibility for the integrity and accuracy of the reporting. Conceptualization: Shuai Yang (SY), David A. Sbarra (DAS), Yann C. Klimentidis (YCK) Methodology: SY, DAS, YCK. Data curation: SY. Software: SY. Formal analysis: SY. Visualization: SY. Writing – original draft: SY. Writing – review & editing: All authors Supervision: DAS, YCK. **Statement on Generative Artificial Intelligence:** Generative AI tools were used to improve language and readability of the paper, and ChatGPT 5.4 was used to assist with statistical code development.

## Abstract

Age-related hearing loss is linked to loneliness and poorer cognitive health, but it remains unclear whether loneliness helps explain associations between hearing difficulties and cognitive performance or dementia, and whether these patterns reflect causal pathways or shared underlying liability. In this preregistered study, we triangulated analyses across multiple data sources spanning approximately 18 years of observational data with 8 sources of molecular genetic information to examine whether loneliness helps explain the association between hearing difficulty and cognitive performance, Alzheimer’s disease dementia, and all-cause dementia, and whether hearing-aid use may buffer this association. In longitudinal parallel-process latent growth curve models (*N* = 10,375) using nine waves of longitudinal data from the Survey of Health, Ageing and Retirement in Europe (SHARE), poorer hearing was associated with greater loneliness, and greater loneliness was associated with poorer cognitive performance, consistent with partial mediation. In contrast, worsening hearing over time was not clearly associated with increasing loneliness over time. Cumulative hearing-aid use did not appear to alter long-term loneliness trajectories, although current hearing-aid use weakened the concurrent association between poorer hearing and greater loneliness. In genetic analyses, we found little evidence that hearing phenotypes or loneliness had clear total or indirect effects on Alzheimer’s disease dementia or all-cause dementia. Analyses accounting for shared genetic liability with neuroticism provided some evidence linking loneliness with poorer cognitive performance, and colocalization analyses further suggested shared genetic architecture across hearing, loneliness, cognition, and neuroticism-related traits. Overall, the findings support a robust cross-domain association between poorer hearing, greater loneliness, and poorer cognitive performance, while suggesting that long-term change and genetic evidence are more consistent with shared liability than with a single causal pathway.

---

Cognitive decline and dementia pose pressing global health challenges, with risk profiles that evolve across the life course and increase markedly with age.^1,2^ Dementia is a leading cause of disability and dependency among older adults worldwide, and the World Health Organization has emphasized the urgency of identifying effective strategies for risk reduction and early intervention.^3^ Accordingly, clarifying the influence of modifiable risk factors that can be targeted before clinically manifest dementia is central to prevention efforts.

Among potentially modifiable risk factors, hearing loss (HL) and loneliness are especially relevant to late-life cognitive health. The 2024 Lancet Commission identifies midlife HL as the leading modifiable risk factor for dementia and highlights social disconnection, including loneliness and social isolation, as an important contributor to dementia risk later in life.^1^ Observational studies generally suggest that HL is associated with accelerated cognitive decline and increased dementia risk, potentially through reduced social engagement, reduced auditory input, and increased cognitive load.^4–6^ A parallel literature links loneliness with poorer cognitive trajectories, Alzheimer’s disease dementia (AD dementia), and all-cause dementia, although loneliness may itself partly reflect related processes such as social disengagement, affective symptoms, stress-related pathways, and diminished cognitive and social resilience.^7–9^ Importantly, longitudinal observational studies remain highly informative for understanding temporal ordering and change, but many prior studies rely on limited follow-up periods and therefore cannot fully distinguish mediation from confounding, or reverse causation. These limitations motivate designs that can better characterize trajectories across time while also strengthening causal inference.

While longitudinal observational studies can establish and replicate associations of interest, genetically-informed methods can help evaluate whether these associations are consistent with causal effects. Mendelian randomization (MR) uses genetic variants as instrumental variables to strengthen causal inference under specific assumptions.^10,11^ Recent MR studies have examined hearing impairment, loneliness, and social isolation in relation to cognitive performance, AD dementia, and all-cause dementia, but results are mixed and the evidence remains incomplete, particularly regarding whether HL and loneliness operate independently or along the same pathway.^12–15^ In the current study, we complete a more integrative, preregistered evaluation of whether HL and loneliness show independent, overlapping, or potentially mediated associations with cognitive outcomes (COs).

A central unresolved question is whether HL contributes to poorer cognitive outcomes partly through loneliness, or whether HL and cognitive decline instead co-occur because of shared underlying processes. HL can constrain communication and social engagement, and observational evidence supports a pathway in which auditory difficulties contribute to social withdrawal and feelings of disconnection;^16,17^ loneliness, in turn, is associated with cognitive decline, AD dementia, and all-cause dementia.^7–9,16^ These findings motivate a mediating hypothesis in which HL indirectly contributes to poorer cognitive health by increasing loneliness. At the same time, HL may also index shared biological processes linked to cognitive aging, including vascular dysfunction, neuroinflammation, and oxidative stress, raising the possibility that HL and cognitive decline co-occur because of common underlying mechanisms and leaving open the potential for reverse effects.^18,19^ If social mechanisms are central, auditory rehabilitation (e.g., hearing aids) might be expected to buffer the association between HL and loneliness, although prior evidence is limited and mixed.^16,20,21^ At the same time, associations between loneliness and cognitive outcomes may not reflect a purely social pathway, because correlated affective and personality-related traits may also contribute to these links. One such trait is neuroticism. Characterized by a tendency toward negative affect, worry, and emotional vulnerability, neuroticism is genetically and phenotypically related to loneliness and also linked to cognitive decline and dementia risk.^7,13,22–24^ Neuroticism may therefore be relevant as a correlated trait that could partly account for associations between loneliness and cognitive outcomes, and may also shape how individuals experience and interpret social disconnection and hearing-related difficulties. A key question, then, is whether loneliness relates to cognitive performance, AD dementia, and all-cause dementia independently of neuroticism, or whether these associations are better understood as reflecting shared liability. Addressing this question requires approaches that can better separate the contributions of correlated traits.

## The Present Study

To build a more causally informative account of how HL, loneliness, and cognitive outcomes are related across the aging spectrum, this paper triangulates evidence across complementary designs through a series of preregistered analyses (https://osf.io/xqdhc/overview). We investigated cognitive outcomes spanning cognitive performance, AD dementia, and all-cause dementia, thereby capturing the continuum of cognitive aging from earlier variation in performance to later clinical impairment.

Cognitive performance may capture earlier differences or decline before the clinical manifestation of dementia, whereas dementia reflects more severe impairment that disrupts daily functioning and independence.^1,25,26^ Considering these outcomes together helps situate potential risk factors across the trajectory of cognitive aging, from earlier changes in performance to later clinical impairment.

Using multi-wave data from the Survey of Health, Ageing and Retirement in Europe (SHARE), we first applied a parallel-process LGCM to test the hypothesis that worsening HL is associated with increases in loneliness and that increases in loneliness are, in turn, associated with steeper declines in cognitive performance; we additionally tested whether hearing-aid use attenuates the HL–loneliness coupling over time.^27,28^ Next, we conducted two-sample MR and MVMR with large-scale Genome Wide Association Studies (GWAS) of HL phenotypes, loneliness, neuroticism, cognitive performance, AD, and dementia to evaluate causal links among HL, loneliness, and cognitive outcomes—including evidence consistent with loneliness mediation—and to test whether loneliness effects persist after accounting for neuroticism.^10,29,30^ Finally, we used colocalization to assess whether associations are driven by shared causal variants or by distinct, LD-linked signals.^31^ Together, these preregistered analyses aim to clarify whether and how HL and loneliness shape cognitive aging across the continuum from cognitive performance to clinical outcomes using the best available methods to address potential confounding, reverse causation, and shared genetic architecture.

## METHODS

### Samples

#### Longitudinal sample

We used data from the Survey of Health, Ageing and Retirement in Europe (SHARE), a cross-national panel study of adults aged 50 years and older that has collected harmonized health, socioeconomic, and social data since 2004 on a roughly biennial schedule. At the time of analysis, SHARE included up to nine waves of data from participants in 28 European countries and Israel.^27^ For the longitudinal analyses, we constructed a de-duplicated person-by-wave panel from harmonized SHARE data and created a person-level wide dataset for structural equation modeling. To ensure that growth parameters were estimable for each process, we required at least two valid observations for hearing across Waves 1, 2, and 4–7, at least two observations for loneliness across Waves 7–9, and at least two observations for objective cognition across Waves 7–9. Hearing at Wave 3 was excluded from the hearing growth indicators because of documented incompatibilities in wording and response format. The final analytic latent growth curve modeling (LGCM) sample included 10,375 participants (mean age = 71.43 years, SD = 7.79; 58.2% women). Sample sizes for key variables across waves are reported in Supplementary Table S1.

#### Genetic samples

For the genetic analyses, we used summary statistics from 8 large-scale genome-wide association studies (GWAS) of hearing-related phenotypes, loneliness, neuroticism, cognitive performance, AD dementia, and all-cause dementia. To reduce bias from population stratification and improve comparability across analyses, the main MR and colocalization analyses were restricted to individuals of predominantly European ancestry. Summary statistics were drawn primarily from the UK Biobank (UKB), FinnGen, and the Million Veteran Program (MVP), together with other published consortia-level datasets described in Table 1.^32–34^

**Table 1.**
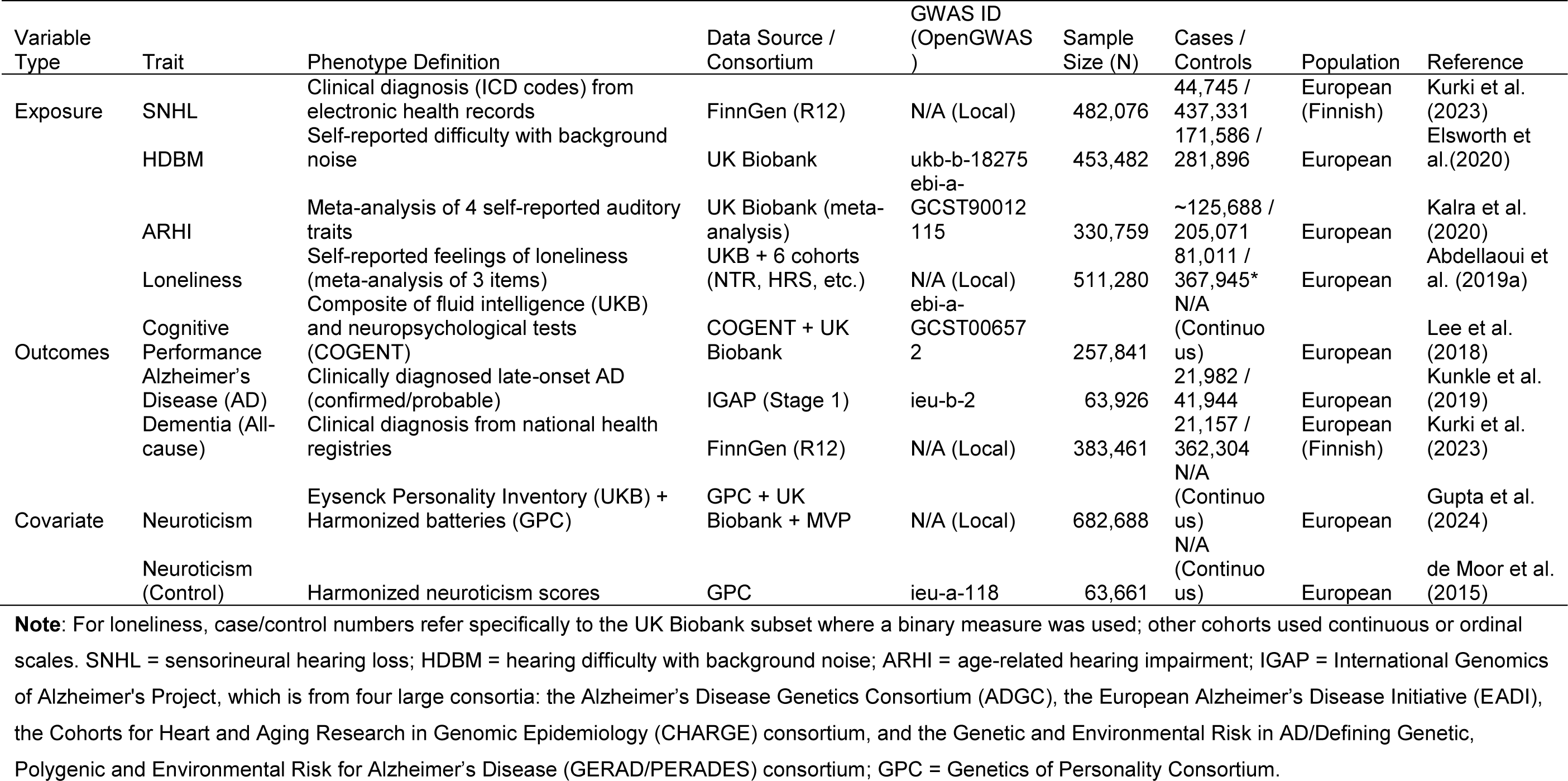
GWAS Used in MR Analyses.

**Table 2.**
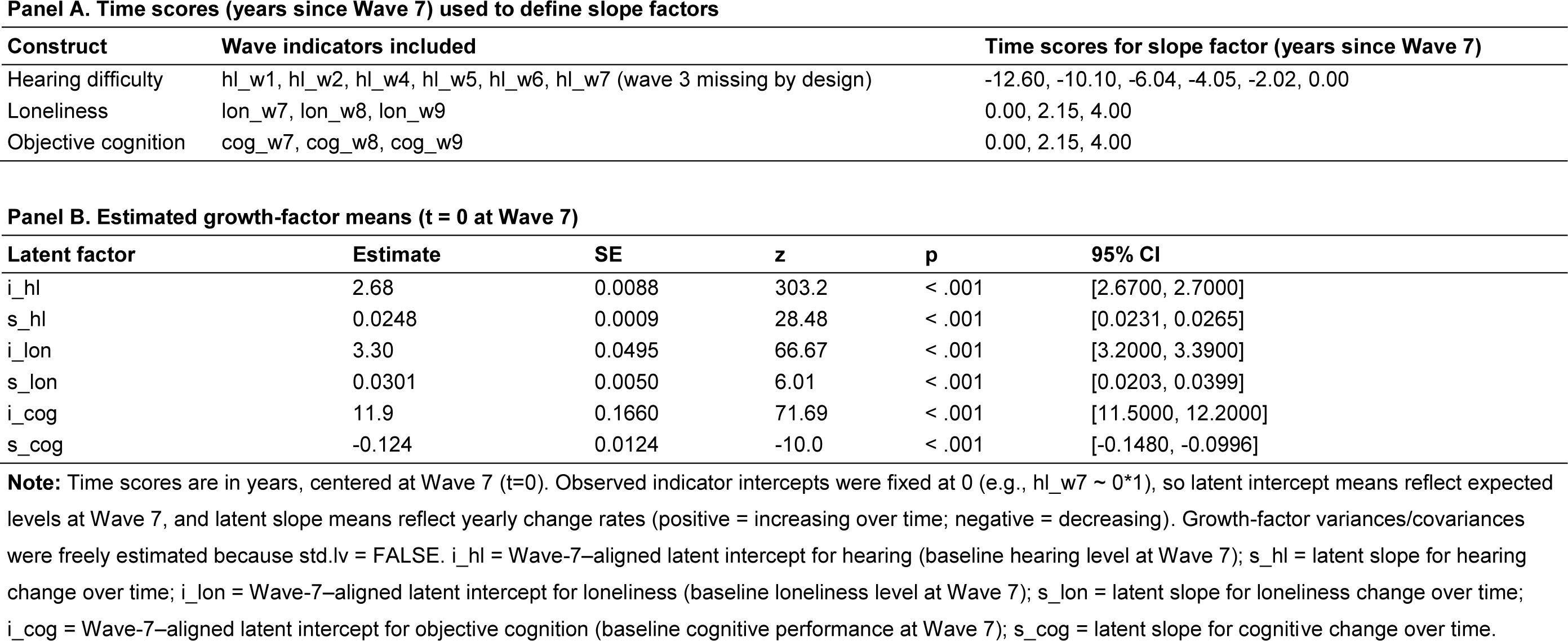
Parallel-process latent growth curve model specification and growth-factor means for hearing difficulty, loneliness, and objective cognition (MLR with FIML; N = 10,375).

Our primary hearing-loss exposure for MR was sensorineural hearing loss (SNHL) diagnosis from FinnGen R12, supplemented by hearing difficulty in background noise (HDBM) and age-related hearing impairment (ARHI) from UKB. Loneliness was instrumented using the largest available meta-analysis.^13^ Cognitive outcomes included cognitive performance, AD dementia, and all-cause dementia. Neuroticism instruments for the primary multivariable MR (MVMR) analyses were derived from a recent large GWAS meta-analysis.^35^ To further reduce concerns about sample overlap, we repeated key neuroticism-adjusted analyses using an independent neuroticism GWAS from the Genetics of Personality Consortium (GPC).

### Measures

#### Hearing

In SHARE, hearing was assessed using the standard self-rated hearing item (1 = excellent, 2 = very good, 3 = good, 4 = fair, 5 = poor), explicitly “taking into account the use of a hearing aid as usually worn.” Self-reported hearing measures are widely used in population-based studies of aging as pragmatic indicators of perceived hearing difficulty, although they do not fully substitute for objective audiometric assessment.^36,37^ We treated this 1–5 rating as continuous, with higher values indicating poorer hearing.

#### Loneliness

Loneliness was measured using the SHARE loneliness score derived from the three-item short form of the Revised UCLA Loneliness Scale, administered in Waves 7–9. The Three-Item Loneliness Scale was developed for large population-based surveys and is implemented in SHARE as a brief measure of perceived social disconnection, with scores ranging from 3 to 9.^38,39^

#### Objective cognition

Objective cognition was operationalized as total word recall (0–20), calculated as the sum of immediate and delayed recall responses to a 10-word list in each wave. Immediate and delayed word recall tasks are widely used in aging research as indicators of episodic memory and cognitive performance.^40^

#### Hearing-aid use

Hearing-aid exposure was operationalized in two ways: Cumulative hearing-aid use through Wave 7 and current hearing-aid use at a given wave. Additional coding details are provided in Supplementary Methods 1.

#### Covariates

Covariates in the longitudinal analyses included age, sex, household income, and neuroticism. To align covariates with the Wave 7 baseline of loneliness and cognition, we prioritized Wave 7 values when available and otherwise used the earliest non-missing value. Sex was coded as binary (0 = male, 1 = female). Household income was based on the SHARE imputed total household income variable and transformed as log(1 + income) to reduce skewness. Neuroticism was assessed using the two-item neuroticism subscale of the BFI-10 and was treated as continuous. Age, transformed income, and neuroticism were z-standardized before modeling. In the longitudinal models, neuroticism was included as a covariate to reduce confounding by stable affective and personality-related differences that may be related to both loneliness and cognitive outcomes.

### Analytic Strategy

All analyses followed our preregistered plan and hypotheses (OSF; https://osf.io/xqdhc/overview).

#### Longitudinal latent growth curve modeling

We used parallel-process LGCMs to test whether changes in loneliness mediated the association between changes in hearing and changes in objective cognition. Hearing was modeled across Waves 1–7, excluding Wave 3 as noted above, and loneliness and objective cognition were modeled across Waves 7–9. This structure was designed to align hearing at Wave 7 with the baseline wave for loneliness and cognition and to impose temporal ordering for the primary mediation analyses.

For each domain, we estimated a latent intercept and a latent linear slope. The primary model evaluated mediation at both the aligned level and the change level. At the aligned level, loneliness was regressed on hearing and covariates, and cognition was regressed on loneliness, hearing, and covariates. At the change level, the loneliness slope was regressed on the hearing slope and covariates, and the cognition slope was regressed on the loneliness slope, the hearing slope, and covariates. Indirect and total effects were defined as the product and sum of the relevant path coefficients, respectively. To evaluate the robustness of slope-based mediation inferences, we fit two prespecified sensitivity models that varied the handling of intercept- and slope-level cross-domain relations. Full model specifications are provided in Supplementary Methods 1. The conceptual diagram of the primary LGCM is presented in Figure 1.

**Figure 1.**
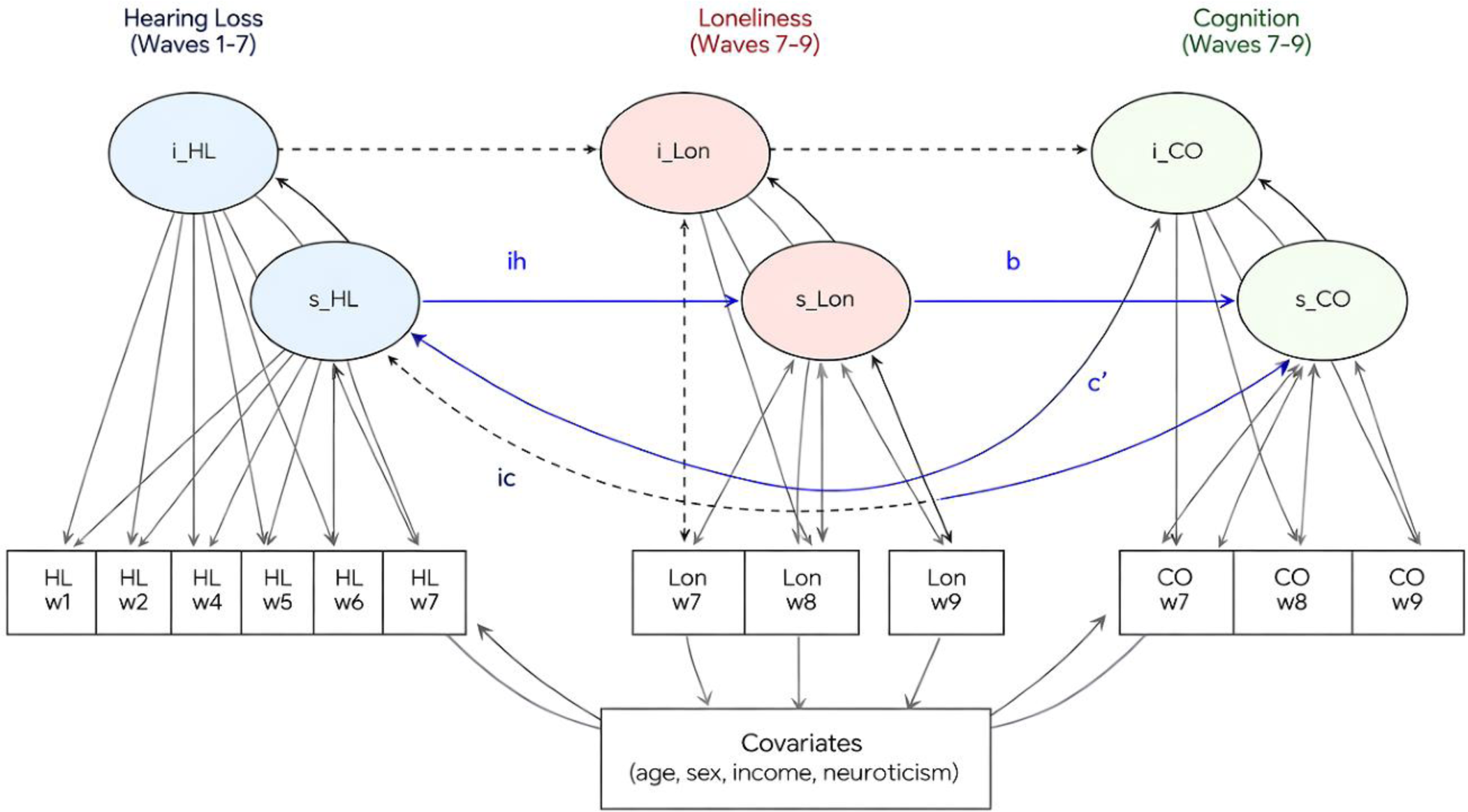
Parallel-process latent growth curve mediation model linking hearing loss (waves 1-7), loneliness (waves 7–9), and cognition (waves 7-9). **Note.** The figure depicts the primary preregistered parallel-process latent growth curve model. Hearing difficulty was modeled across SHARE Waves 1, 2, and 4–7, with Wave 3 excluded because of incompatibilities in wording and response format; loneliness and objective cognition were modeled across Waves 7–9. For each domain, a latent intercept and a latent linear slope were estimated, with time centered at Wave 7 so that intercepts represent Wave-7-aligned levels and slopes represent change per year. The model evaluated mediation at both the aligned level and the change level: loneliness was regressed on hearing and covariates, and cognition was regressed on loneliness, hearing, and covariates at the intercept level; the loneliness slope was regressed on the hearing slope and covariates, and the cognition slope was regressed on the loneliness slope, the hearing slope, and covariates at the change level. Covariates included age, sex, household income, and neuroticism.

**Figure 2.**
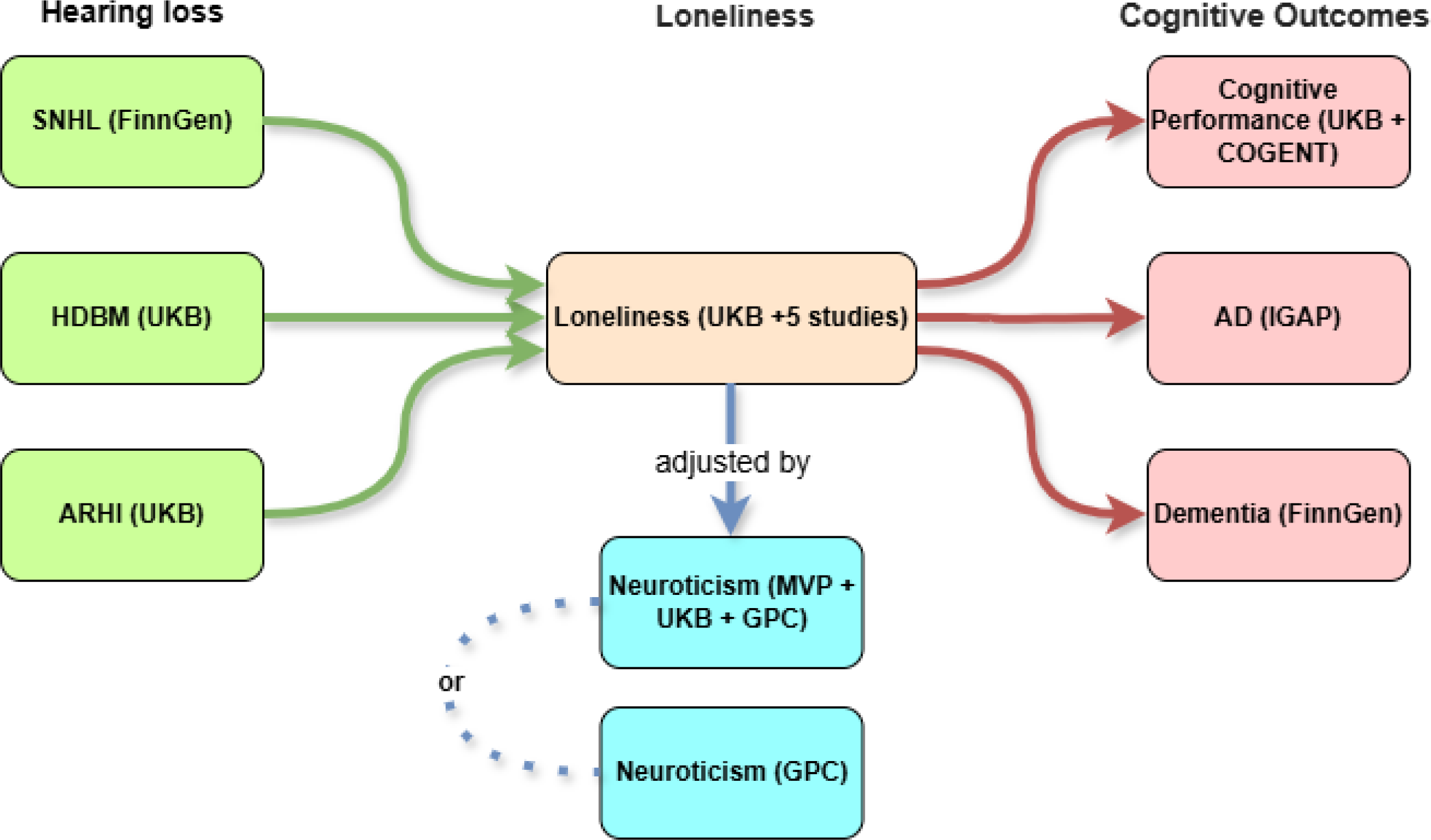
Overall analytic workflow of MR analyses.

All LGCMs were estimated in R (version 4.5.2) using the lavaan package (Rosseel, 2012) with maximum likelihood and robust corrections (MLR).^41^ Missing outcome data were handled using full-information maximum likelihood under a missing-at-random assumption. Model fit was evaluated using the comparative fit index (CFI), Tucker–Lewis index (TLI), root mean square error of approximation (RMSEA), and standardized root mean square residual (SRMR).

#### Hearing-aid moderation analyses

Because hearing aids are a clinically relevant management strategy for hearing loss, we conducted four complementary moderation analyses to test whether hearing-aid use altered the hearing–loneliness association. These analyses examined both cumulative hearing-aid exposure through Wave 7 and current hearing-aid use at a given wave and included adjustment for age, sex, household income, and neuroticism. Briefly, we evaluated moderation using (1) factor-score-based models of loneliness change, (2) mixed-effects models of loneliness trajectories from Waves 7–9, (3) Wave 7 cross-sectional models, and (4) all-wave mixed-effects models of occasion-specific hearing and loneliness. Full coding rules and model specifications are provided in Supplementary Methods 1.

#### Mendelian randomization analyses

We used two-sample MR to evaluate whether genetically predicted hearing-related phenotypes were associated with loneliness and cognitive outcomes. MR estimates can be interpreted causally only under the standard instrumental-variable assumptions. We used MVMR to evaluate whether associations involving loneliness persisted after accounting for genetic liability to neuroticism, and we used a two-step MR framework to assess whether loneliness showed evidence consistent with mediation in the pathway from hearing phenotypes to cognitive outcomes.

For all exposure traits, we selected genome-wide significant variants (P < 5 × 10⁻□) and clumped them for independence using an LD threshold of r² < 0.001 within a 10,000 kb window based on the 1000 Genomes European reference panel (The 1000 Genomes Project Consortium, 2015). We harmonized exposure and outcome summary statistics to align effect alleles and excluded palindromic variants with intermediate allele frequencies or incompatible alleles. Instrument strength was assessed using F-statistics and summarized in the Supplement as a diagnostic of potential weak-instrument bias.

Primary MR analyses estimated the total effects of four exposures (SNHL, HDBM, ARHI, and loneliness) on three outcomes (cognitive performance, AD dementia, and all-cause dementia), yielding 12 primary exposure–outcome pathways. The inverse-variance weighted (IVW) estimator was used as the primary method. For the primary hypothesis set, we used a Bonferroni-corrected significance threshold of P < .004 (0.05/12). Uncorrected P values are reported descriptively, and false discovery rate (FDR)-adjusted results are provided in the Supplement for completeness.

For two-step MR mediation analyses, we estimated indirect effects as the product of the exposure-to-mediator effect and the mediator-to-outcome effect, with standard errors and 95% confidence intervals derived using the delta method. To reduce bias from sample overlap, we prioritized three mediation pathways: SNHL → loneliness → cognitive performance, SNHL → loneliness → AD dementia, and HDBM or ARHI → loneliness → all-cause dementia.

For MVMR, we fit separate models in which each hearing phenotype was modeled jointly with loneliness in relation to cognitive outcomes, and we then conducted additional models that included neuroticism. Our primary MVMR specification used a union-of-instruments approach, in which genome-wide significant variants from all modeled exposures were pooled, re-clumped for independence, and harmonized across exposures and outcomes. Full details on alternative instrument-construction strategies and sensitivity analyses are provided in Supplementary Methods 1.

We conducted complementary sensitivity analyses using the weighted median estimator, the MR-Egger intercept test, Cochran’s Q statistic, and MR-PRESSO. All MR analyses were performed in R (version 4.5.2) using TwoSampleMR, MR-PRESSO, and ieugwasr.^42–44^

### Colocalization analyses

We conducted Bayesian colocalization analyses to evaluate whether trait pairs shared a regional causal signal within the same genomic locus. These analyses were used to complement MR by clarifying whether pairs of traits were likely to reflect a shared regional signal rather than clearly distinct local association signals. Colocalization does not on its own distinguish mediation from horizontal pleiotropy, but it is informative for interpreting whether MR findings may reflect shared local genetic architecture.

We analyzed a prespecified set of 22 trait pairs spanning key hearing, loneliness, neuroticism, and cognitive-outcome relationships. Using full GWAS summary statistics for each trait, we identified approximately independent lead loci, extracted ±500 kb windows around each locus, and proceeded when at least 50 variants overlapped between the two datasets within the region. Analyses were performed in R (version 4.5.2) using the coloc package (coloc.abf), which estimates posterior probabilities for five competing hypotheses within a locus.^31^ We interpreted PP₄ ≥ 0.80 as strong evidence of colocalization and PP₃ > 0.50 as evidence consistent with distinct causal variants within the same region. Full implementation details, including priors and harmonization procedures, are provided in Supplementary Methods 2.

## RESULTS

### Results—LGCM

#### Longitudinal change in hearing, loneliness, and objective cognition

We first characterized average change and individual differences in self-rated hearing, loneliness, and objective cognition across the SHARE waves used in the preregistered LGCM component. The parallel-process LGCM (with the hearing intercept centered at Wave 7 and slopes expressed in raw units per year) fit the data well (robust/scaled χ²(89) = 1450.0, p < .001; CFI = 0.964; TLI = 0.954; RMSEA = 0.0385; SRMR = 0.064). Estimated latent means indicated that, on average, self-rated hearing worsened over approximately 12.6 years of modeled SHARE hearing assessments, as the mean age of participants increased from 59.9 to 71.5 years (Wave-7–aligned hearing intercept: *i*HL = 2.68; hearing slope: *s*HL = 0.0248 units/year). Loneliness also increased over follow-up (*i*Lon = 3.30; *s*Lon = 0.0301 units/year), whereas objective cognition declined (*i*CO = 11.9; *s*CO = −0.124 units/year). Variance components (Table 2) indicated meaningful between-person heterogeneity in trajectories, supporting a parallel-process approach rather than relying on cross-sectional associations alone. Figure 3 visualizes the estimated mean trajectories and uncertainty bands.

**Figure 3.**
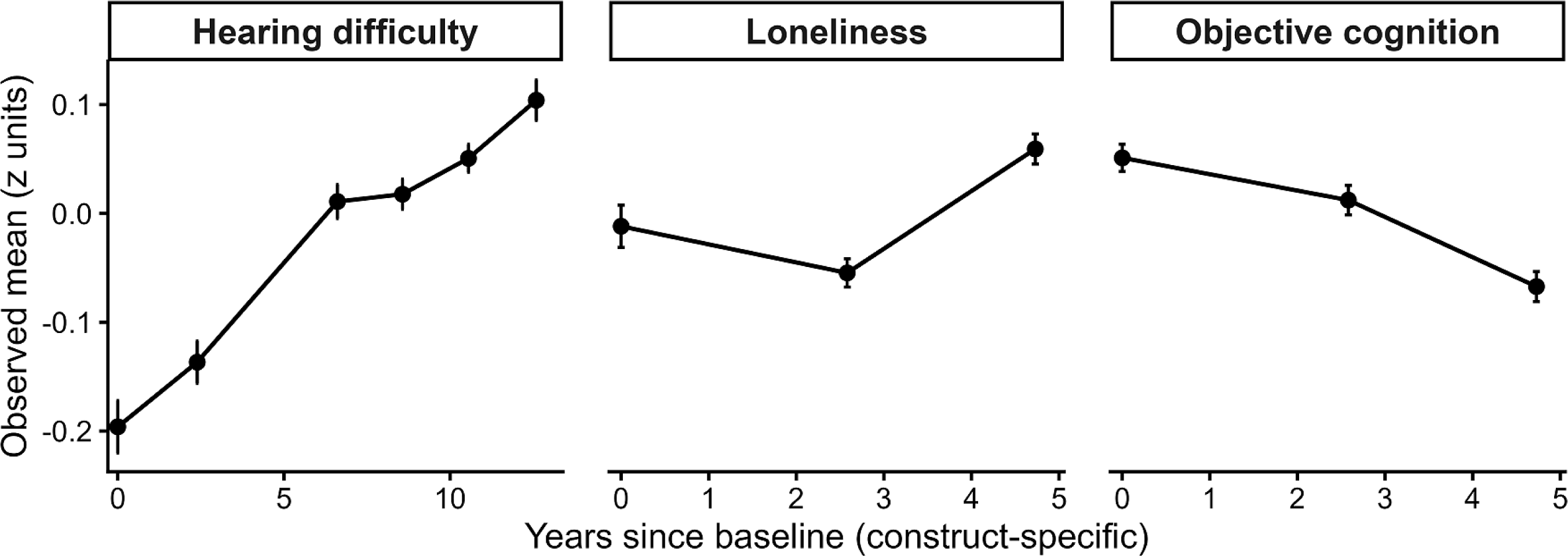
Mean trajectories of hearing difficulty, loneliness, and objective cognition in the LGCM analytic sample (N = 10,375). **Note:** Points represent observed means and error bars indicate 95% confidence intervals. Values are expressed in z-standardized units within the LGCM analytic sample for visualization only (LGCM estimation used raw indicator units). The x-axis is construct-specific, with time scaled to years since the first available measurement for each construct (hearing: Waves 1-7, with Wave 3 omitted by design; loneliness and cognition: Waves 7–9). Higher scores indicate worse hearing difficulty and greater loneliness; higher cognition scores indicate better performance.

#### Preregistered hypotheses: coupling and longitudinal mediation

We next tested preregistered hypotheses concerning longitudinal coupling and mediation in the SHARE LGCMs. Our primary preregistered hypothesis focused on change processes: worsening hearing over time would be associated with increasing loneliness over time, which in turn would be associated with steeper declines in objective cognitive performance. Wave-7–aligned level paths were estimated in the same model to describe cross-domain associations at the aligned baseline level, but these intercept-level paths were not part of the primary preregistered hypotheses.

#### Level (Wave 7 aligned)

Poorer Wave-7–aligned hearing was associated with higher Wave-7 loneliness (*i*Lon on *i*HL: β = 0.135, SE = 0.017, p < .001). In turn, higher Wave-7 loneliness was associated with lower Wave-7 objective cognition (*i*CO on *i*Lon: β = −0.468, SE = 0.034, p < .001). A direct association between Wave-7 hearing and Wave-7 cognition also remained (*i*CO on *i*HL: β = −0.353, SE = 0.042, p < .001). The resulting indirect level effect was statistically significant (β_indirect(level) = −0.0631, SE = 0.0092, p < .001), and the total level effect was β_total(level) = −0.416, SE = 0.0426, p < .001, consistent with partial mediation at the Wave-7–aligned level (Table 3).

**Table 3.**
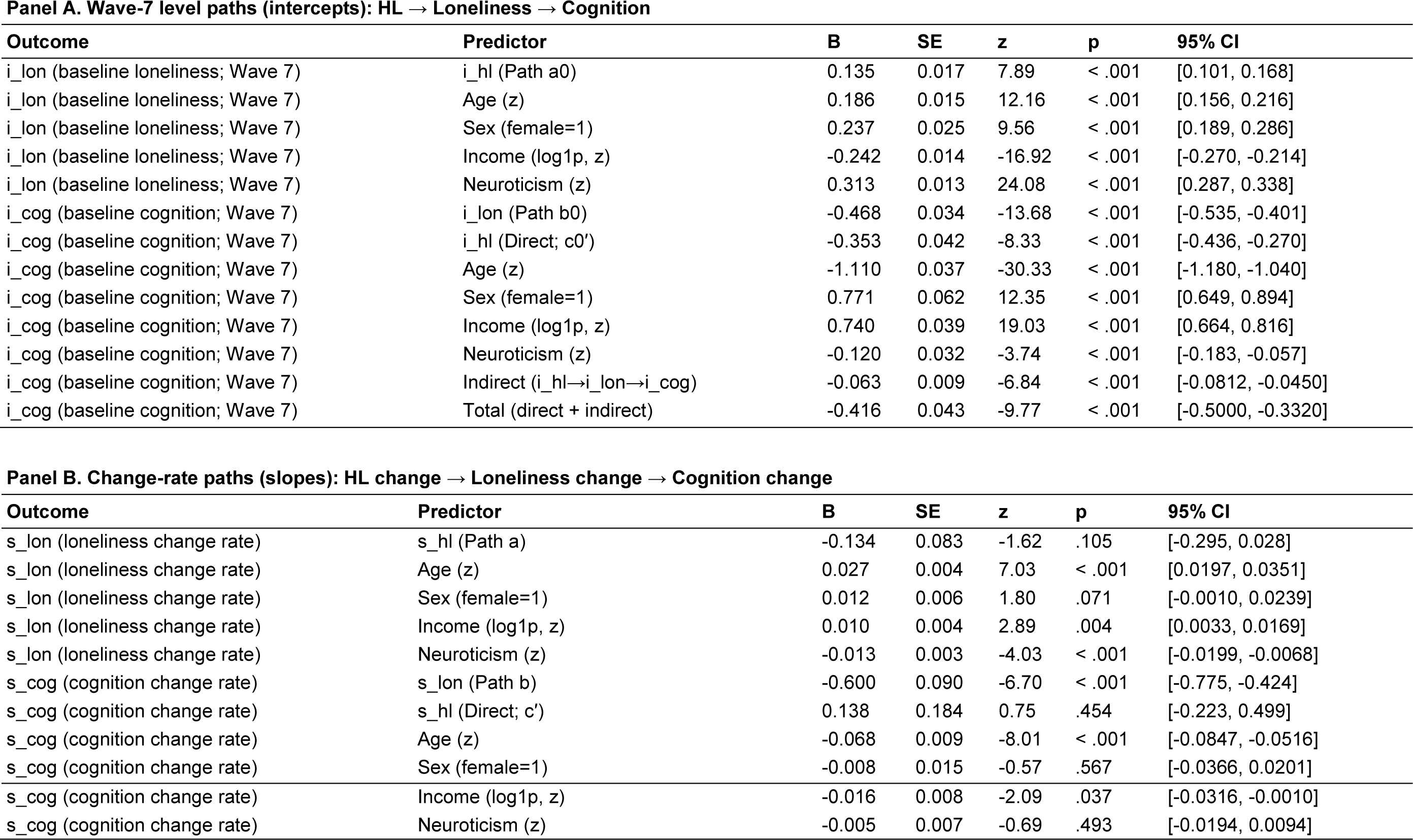
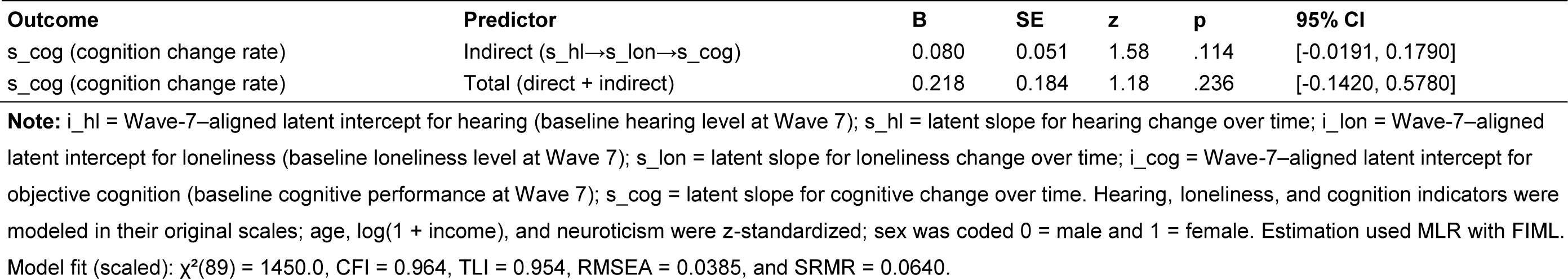
Preregistered mediation and covariate effects on change parameters (and Wave-7 level parameters), parallel-process LGCM (MLR, FIML; N = 10,375).

#### Change (slopes; raw units/year)

In the fully adjusted primary model, the preregistered slope-to-slope coupling between hearing change and loneliness change was not statistically significant (*s*Lon on *s*HL: β = −0.134, SE = 0.083, p = .105). However, increases in loneliness over time were strongly associated with declines in objective cognition (*s*CO on *s*Lon: β = −0.600, SE = 0.090, p < .001). The direct slope path from hearing change to cognitive change was not significant (*s*CO on *s*HL: β = 0.138, SE = 0.184, p = .454). Consequently, the indirect slope effect was not statistically significant in the primary model (β_indirect(slope) = 0.0802, SE = 0.0507, p = .114), nor was the total slope effect (β_total(slope) = 0.218, SE = 0.184, p = .236). In a prespecified slope-only sensitivity model, the indirect slope effect was statistically significant (β_indirect(slope) = 0.151, SE = 0.0511, p = .003), whereas the level-only sensitivity model did not converge.

#### Covariates and the stability of the longitudinal pathways

At the Wave-7–aligned level, covariates showed clear patterning consistent with established aging correlates. Wave-7–aligned loneliness levels were higher among older participants (β = 0.186, p < .001), females (β = 0.237, p < .001), those with lower income (β = −0.242, p < .001), and those higher in neuroticism (β = 0.313, p < .001). Wave-7 cognition levels were lower with older age (β = −1.11, p < .001) and higher with female sex (β = 0.771, p < .001) and higher income (β = 0.740, p < .001); neuroticism was negatively associated with Wave-7 cognition (β = −0.120, p < .001).

For change processes, older age predicted faster increases in loneliness (*s*Lon: β = 0.0274, p < .001) and faster cognitive decline (*s*CO: β = −0.0681, p < .001). Income was associated with loneliness change (β = 0.0101, p = .0036) and cognitive change (β = −0.0163, p = .0367) in the fully adjusted slope equations. Neuroticism was associated with loneliness change (β = −0.0133, p < .001) but was not a significant predictor of cognitive change (β = −0.0050, p = .493) in the primary model’s slope structure. Table 3 reports the preregistered focal paths and derived indirect/total effects from the fully adjusted model.

#### Hearing-aid use as a preregistered moderator of the hearing–loneliness link (A–D)

We conducted four moderation analyses to test whether hearing-aid use buffered the hearing–loneliness association.

***A) Latent slope moderation (factor-score approach).*** In a regression model including the hearing slope factor score, cumulative hearing-aid use, and their interaction, the interaction term was not significant (β = 0.172, SE = 0.144, p = .232; N = 10,375). This finding provided no evidence that cumulative hearing-aid use moderated the association between hearing change and loneliness change in this specification.
***B) Time × Cumulative hearing-aid use predicting loneliness trajectory (Waves 7–9).*** In mixed-effects models of standardized loneliness across Waves 7–9, the time × Cumulative hearing-aid use interaction was not significant (β = 0.00292, SE = 0.0125, |t| = 0.234, p = .815; 30,339 observations from 13,550 participants), providing convergent evidence that Cumulative hearing-aid use did not alter the average rate of loneliness change over the Wave-7–anchored follow-up window.
***C) Wave-7 cross-sectional moderation (HL × current HA).*** At Wave 7, poorer hearing was associated with higher loneliness (β = 0.0833, SE = 0.0087, p < .001; N = 13,263), but the HL × HA interaction was not significant (β = −0.0372, SE = 0.0297, p = .210), indicating no detectable moderation at Wave 7 in the cross-sectional model.
***D) All-wave occasion-specific moderation (mixed models; earliest-baseline covariates).*** In mixed-effects models using all available observations with non-missing hearing, current hearing-aid use, and loneliness (94,253 observations from 44,004 participants), poorer hearing was strongly associated with higher concurrent loneliness (β = 0.0794, SE = 0.0035, |t| = 22.79). This association was significantly attenuated among hearing-aid users (HL × HA: β = −0.0289, SE = 0.0124, |t| = −2.34, p = .019). Including wave fixed effects produced a similar interaction estimate (β = −0.0259, SE = 0.0124, |t| = −2.09, p = .036). In within/between decomposition models, moderation was concentrated in the within-person component (HA × within-person HL deviation: β = −0.0454, SE = 0.0163, |t| = −2.79, p = .005), whereas the between-person component interaction was not significant (β = 0.0183, SE = 0.0188, |t| = 0.97).

## Results—Mendelian randomization

### Primary (preregistered) causal tests: HL and loneliness on COs

Consistent with our preregistered plan, we first tested univariable MR associations of loneliness and each hearing phenotype (SNHL, HDBM, and ARHI) with three cognitive outcomes (cognitive performance, AD, and all-cause dementia), yielding 12 exposure–outcome combinations. Across these tests, IVW estimates were uniformly small, imprecise, and not statistically significant. For example, the IVW effect of loneliness on cognitive performance was β = 0.018 (SE = 0.166, p = .91), and the effect of SNHL on cognitive performance was β = 0.019 (SE = 0.016, p = .23). None of the primary IVW estimates survived Bonferroni correction or corresponding FDR control.

### Preregistered two-step MR mediation: HL → loneliness → COs

Across the prespecified mediation pathways (e.g., SNHL → Loneliness → Cognitive performance; SNHL → Loneliness → AD; HDBM/ARHI → Loneliness → Dementia), the estimated mediated proportions were small and non-significant, with wide confidence intervals spanning zero. Thus, within the current MR setting, we found no clear evidence that loneliness carries a substantial proportion of the genetic effect of hearing loss on cognitive performance, AD, or dementia.

### Preregistered MVMR (mutual adjustment): independent effects of hearing loss and loneliness on cognitive outcomes

To disentangle potential independent contributions of hearing loss and loneliness, we implemented MVMR models using a union-of-instruments specification (variants combined across exposures and re-clumped to form a shared, independent instrument set). When SNHL and loneliness were modeled jointly as predictors of dementia or cognitive performance, neither trait showed evidence of a direct effect after mutual adjustment. For example, in the SNHL–loneliness model for dementia, the adjusted effect of loneliness was β = −0.17 (SE = 0.20, p = .39), and the adjusted effect of SNHL was β = 0.05 (SE = 0.05, p = .30). Parallel MVMR analyses for HDBM and ARHI (each paired with loneliness) yielded similarly null and imprecise estimates across cognitive performance, AD, and dementia. Collectively, these preregistered MVMR results do not indicate strong, independent genetic effects of either hearing loss or loneliness on the cognitive outcomes examined.

### Genetic analyses involving neuroticism

Because neuroticism is strongly genetically correlated with loneliness and may confound or share liability with cognitive outcomes, we conducted MR analyses focusing on neuroticism–loneliness coupling and neuroticism-related pathways. In univariable MR, we observed strong bidirectional associations between neuroticism and loneliness: genetically proxied neuroticism predicted higher loneliness (β = 0.57, SE = 0.024, p = 6.6 × 10⁻¹²), and genetically proxied loneliness predicted higher neuroticism (β = 0.45, SE = 0.044, p = 1.5 × 10⁻²). Genetically proxied neuroticism was also robustly associated with poorer cognitive performance (β = −0.24, SE = 0.040, p = 1.4 × 10⁻), whereas its effects on AD and all-cause dementia were small and non-significant.

We then used MVMR to test whether the association between loneliness and cognitive performance persisted after accounting for neuroticism. In models using the large meta-analytic neuroticism GWAS, loneliness remained significantly associated with cognitive performance after adjustment for neuroticism (β = −0.23, SE = 0.064, p = 3.7 × 10⁻), whereas the adjusted neuroticism effect was near zero (β = 0.00, SE = 0.044, p = .97). However, when the same analyses were repeated using an independent, smaller neuroticism GWAS from the Genetics of Personality Consortium, neither loneliness nor neuroticism was significantly associated with cognitive performance or dementia (all p ≥ .26).

Finally, we explored joint effects of hearing phenotypes and neuroticism on loneliness with MVMR. In models including HDBM and neuroticism, HDBM was positively associated with loneliness after adjustment (β = 1.03, SE = 0.20, p = 3.7 × 10⁻), whereas in models including ARHI and neuroticism, ARHI showed an association in the opposite direction (β = −0.41, SE = 0.11, p = 2.7 × 10⁻). Given that hearing phenotypes are correlated and differ in measurement and instrument composition, this sign discordance is most consistent with sensitivity of MVMR estimates to instrument choice and residual collinearity rather than truly opposing biology. Notably, these hearing–neuroticism MVMR signals did not persist when using an independent, smaller neuroticism GWAS from GPC (all p ≥ .09), reinforcing that these findings are generally not supportive of a causal effect, at least with existing genetic instruments. Consistent with the primary mediation analyses, neuroticism-adjusted two-step MR mediation estimates (hearing phenotype → loneliness [adjusted] → cognitive outcomes) were small and non-significant, with confidence intervals spanning both negative and positive values.

Full MR results—including all primary IVW estimates, mediation outputs, union-of-instruments MVMR models, neuroticism-based sensitivity analyses, and robustness checks—are reported in Supplementary Tables S2–S12.

## Results—Colocalization

Using our preregistered genome-wide colocalization pipeline, we evaluated evidence that trait pairs share a single causal variant within a region (PP₄ from coloc.abf). We focused on regions that passed harmonization and overlap thresholds (≥50 shared SNPs), and we interpreted PP₄ ≥ 0.80 under the default prior (p₁ = p₂ = 1×10⁻; p₁₂ = 1×10⁻) as strong evidence of colocalization. Across all prespecified trait pairs, we identified 47 independent lead regions with PP₄ ≥ 0.80 (PP₄ range: 0.80–1.00) (Figure 4 and Supplementary Table S13). Importantly, the distribution of these signals was highly uneven across trait pairs, with the majority involving neuroticism in combination with loneliness, hearing phenotypes, or cognitive performance, consistent with neuroticism acting as a pleiotropic hub.

**Figure 4.**
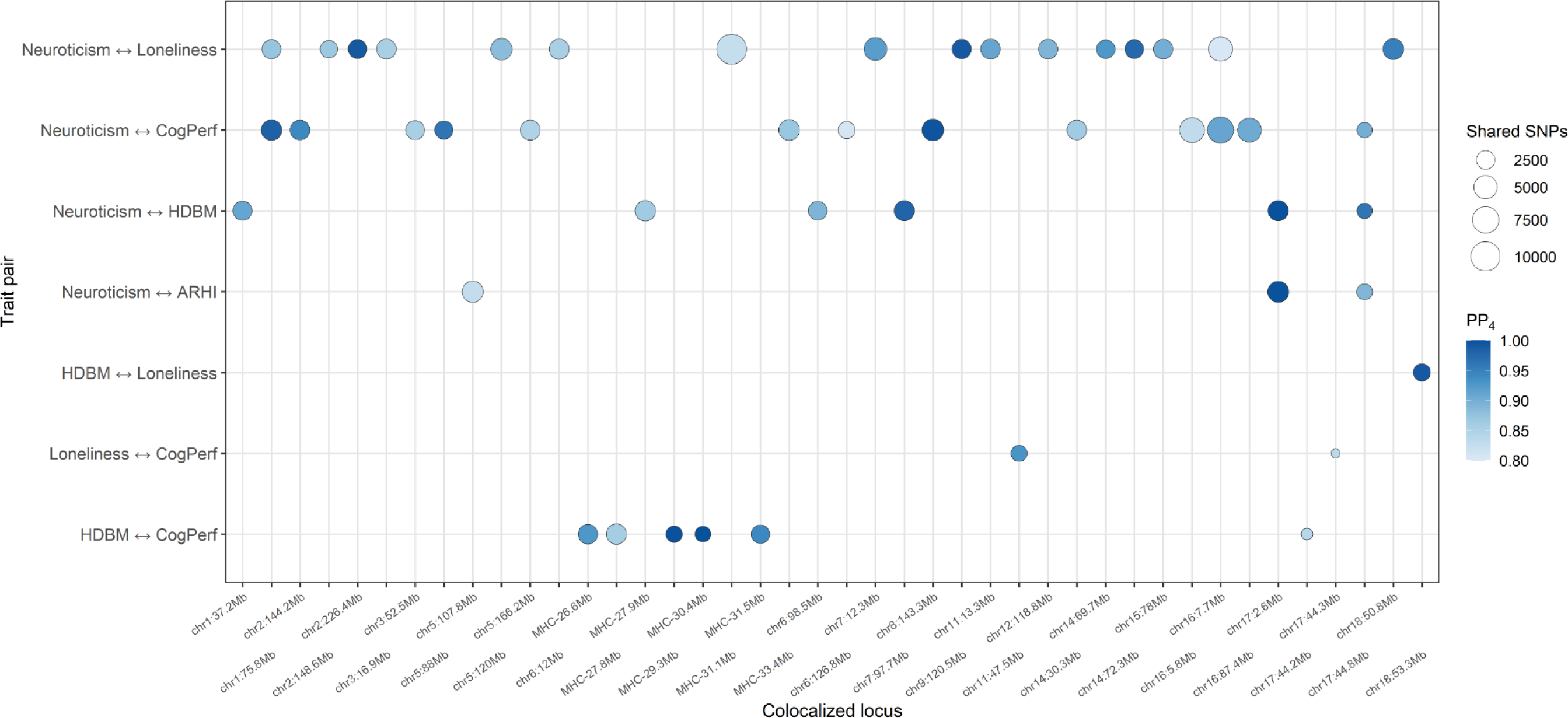
Figure 4. Colocalization architecture across hearing, loneliness, neuroticism, and cognitive traits. **Note:** PP4 denotes the posterior probability for hypothesis 4 in the coloc analysis, indicating support for the two traits sharing a single causal variant within the tested region. Only loci with PP4 ≥ 0.80 under the default prior are shown. The x-axis includes all unique genomic loci with PP4 ≥ 0.80 identified across the prespecified trait pairs. Bubble size represents the number of overlapping SNPs used in the regional colocalization analysis.

Robust colocalization was observed for HDBM and loneliness at a locus on chromosome 18 (lead position chr18:53,252,388; PP₄ = 0.99). Loneliness and cognitive performance colocalized at two loci (chr11:47,523,214, PP₄ = 0.93; chr17:44,344,988, PP₄ = 0.84). The strongest hearing–cognition pattern involved HDBM and cognitive performance in the extended MHC region on chromosome 6, where five clustered loci spanning approximately 26.5–31.5 Mb showed PP₄ ≥ 0.86, with the top signal at chr6:30,441,303 reaching PP₄ = 1.00; an additional locus was observed on chromosome 17 (chr17:44,221,836; PP₄ = 0.84). Neuroticism showed the broadest colocalization footprint, including multiple loci shared with HDBM, ARHI, loneliness, and cognitive performance (Figure 4). For several pairs involving FinnGen dementia or AD GWAS, genomic coverage was reduced by build differences and SNP-alignment requirements, resulting in fewer analyzable regions after quality control.

## DISCUSSION

Three main conclusions emerged from our triangulated analyses. First, in SHARE, poorer hearing at the Wave 7–aligned level was associated with greater loneliness, and greater loneliness was associated with poorer cognitive performance, whereas evidence for change-to-change mediation over time was weaker and sensitive to model specification. Second, cumulative hearing-aid exposure did not moderate long-term change in loneliness, but current hearing-aid use attenuated the concurrent association between poorer hearing and greater loneliness in repeated-measures models. Third, MR analyses provided limited evidence that hearing phenotypes or loneliness had clear causal effects on dementia outcomes with currently available instruments, whereas neuroticism emerged as a central component of the shared genetic architecture linking loneliness and cognitive performance.

### Longitudinal coupling of hearing, loneliness, and cognition in SHARE

Consistent with prior observational work linking hearing difficulty and social disconnection and with broader dementia risk frameworks, SHARE parallel-process LGCMs indicated reliable coupling among Wave-7–aligned level processes.^1,16^ In the primary specification, poorer hearing at Wave 7 was associated with higher loneliness, and higher loneliness was associated with lower objective cognitive performance, yielding a statistically reliable indirect effect through loneliness alongside a remaining direct hearing–cognition association. These findings indicate that loneliness accounted for part, but not all, of the cross-domain association between hearing and cognition at the Wave-7–aligned level.

In contrast, evidence for slope-level (change-to-change) mediation was weaker in the fully adjusted primary model, with the indirect slope effect not statistically distinguishable from zero. In a prespecified slope-only sensitivity model, the indirect slope effect was statistically significant, whereas the level-only sensitivity model did not converge. Taken together, the SHARE findings provide stronger support for Wave-7–aligned level associations than for coordinated change over time.

### Hearing aids: no moderation of long-term slopes, but buffering of concurrent burden

Hearing rehabilitation can reduce communication burden and improve social functioning, although effects on loneliness and social isolation appear modest and heterogeneous across studies. Recent trial and review evidence suggests that hearing interventions may preserve social connectedness and improve some loneliness-related outcomes, but prior observational and longitudinal studies have not consistently shown durable changes in loneliness over time.^21,45–48^

At the trajectory level, we found no evidence that cumulative hearing-aid exposure moderated coupling between hearing-change and loneliness-change (latent slope interaction), and no evidence that cumulative hearing-aid exposure altered average within-person change in loneliness over years since Wave 7 in time-based mixed models. Thus, within the overall follow-up window and measurement schedule available, hearing-aid exposure was not associated with a systematically weaker rate of loneliness increase given hearing decline in these long-term change-focused tests.

At the occasion level, however, all-wave mixed-effects models indicated that current hearing-aid use attenuated the concurrent association between poorer hearing and higher loneliness.

Decomposition of hearing into between-person and within-person components further suggested that this moderation was driven primarily by within-person fluctuations (i.e., deviations from an individual’s typical hearing level), rather than stable between-person differences. This within-person moderation is both substantively meaningful (a potential buffering of “acute” burden) and challenging to interpret given multi-year spacing between waves; here, “deviations” reflect differences across years rather than day-to-day fluctuations. Taken together, the results suggest hearing aids may reduce the immediate social–emotional burden of hearing difficulty without necessarily reshaping longer-term loneliness trajectories in this observational setting.

### MR/MVMR: limited evidence for large direct causal effects of HL or loneliness on dementia and global cognition

Across preregistered MR analyses evaluating loneliness and multiple hearing phenotypes on cognitive performance, AD dementia, and all-cause dementia, IVW estimates were generally small and statistically non-significant after correction for multiple testing, and two-step MR mediation analyses yielded small, imprecise mediated proportions. MVMR models that jointly included hearing phenotypes and loneliness (and, in additional models, neuroticism) likewise did not provide consistent evidence that either hearing phenotypes or loneliness exert robust independent effects on dementia outcomes under currently available instruments. These results should not necessarily be interpreted as disconfirming the SHARE associations. Rather, they highlight a familiar tension across designs: MR estimates reflect lifelong genetic liability, whereas SHARE captures late-life phenotypes shaped by environmental exposures, clinical care, and changing social contexts.^6^ In addition, hearing phenotypes differ across GWAS in measurement and coding; therefore, careful harmonization of effect directions and construct alignment is essential when interpreting sign differences across instruments.

### Neuroticism as a key genetic correlate—and the sensitivity of “loneliness-independent” effects

A consistent signal across our genetic analyses was strong coupling between neuroticism and loneliness, aligning with prior molecular genetic evidence that associations between loneliness and personality are largely driven by shared genetic liability with neuroticism.^13^ In our univariable MR analyses, neuroticism showed robust associations with loneliness and with cognitive performance, whereas effects on AD and all-cause dementia were small and non-significant. This pattern reinforces the rationale for treating neuroticism as a central correlated trait when probing loneliness–cognition links in genetic designs.

In MVMR, we evaluated whether loneliness shows evidence of an association with cognitive performance after accounting for neuroticism. Importantly, this inference was instrument-dependent: models using a large meta-analytic neuroticism GWAS suggested a loneliness association with cognitive performance after adjustment, whereas the same conclusion did not replicate when using an alternative, smaller neuroticism GWAS. When exposures are highly genetically correlated, apparent “independent” effects can be sensitive to instrument choice, phenotype definition, and differences in overlap or measurement across GWAS; thus, any loneliness-specific effect on cognitive performance after neuroticism adjustment should be treated as suggestive until replicated with harmonized instruments.

### Colocalization: shared loci concentrated around neuroticism (and select hearing–cognition regions), but no single “mediation locus”

Our genome-wide colocalization results complemented the MR results by distinguishing shared causal variants from distinct signals in linkage disequilibrium. The strongest and most consistent shared-variant signals involved neuroticism, loneliness, and cognitive performance, reinforcing neuroticism as a pleiotropic component of shared architecture.^13^ We also observed select regions suggestive of overlap involving hearing phenotypes and cognitive performance, including signals in highly pleiotropic genomic regions.

At the same time, colocalization did not reveal a single, clean locus that would neatly instantiate a unitary hearing → loneliness → cognition mediation pathway. Rather, the shared architecture was concentrated in neuroticism-related regions, together with a smaller number of hearing–cognition and hearing–loneliness loci. This pattern helps contextualize why MR estimates for a specific mediation pathway can be small and unstable: if underlying biology is polygenic and shared across traits, isolating a strong, exposure-specific causal effect for any single pathway is difficult even with large samples.

### Integrating evidence across methods and implications for prevention

Integrating across methods, our results support a nuanced view of modifiability. Longitudinal evidence is consistent with the idea that hearing difficulty and loneliness are meaningfully linked in later life and that loneliness partially explains the hearing–cognition association at the aligned level, consistent with psychosocial and mechanistic hypotheses in the broader literature.^1,6^ In contrast, genetic analyses did not support large, direct causal effects of hearing phenotypes or loneliness on dementia outcomes with current instruments and instead highlighted neuroticism as a central component of shared genetic liability.

From a clinical and public-health standpoint, the hearing-aid moderation results suggest that amplification may reduce the immediate loneliness burden associated with hearing difficulty, even if it does not measurably shift longer-term loneliness trajectories in these observational analyses. This pattern motivates combined approaches that integrate hearing care with strategies to strengthen social engagement and address emotional vulnerability (including neuroticism-linked processes), while continuing to evaluate downstream cognitive outcomes in designs with denser measurement and intervention follow-up.

### Limitations and strengths

Several limitations warrant caution. SHARE hearing and loneliness are self-reported and may be influenced by reporting biases related to mood and personality; although we modeled longitudinal structure and adjusted for neuroticism and sociodemographic covariates, residual confounding remains possible. Generalizability is limited by the predominantly European-ancestry composition of SHARE and the GWAS datasets. MR relies on instrumental-variable assumptions and adequate instrument strength, and dementia outcomes are heterogeneous with power constraints. Offsetting these limitations, strengths include the preregistered confirmatory framework, the multi-wave SHARE longitudinal design with temporal ordering (hearing preceding loneliness/cognition measurement windows), and integration of complementary genetic approaches to probe causality and shared etiologic architecture.

### Conclusion

In conclusion, the preregistered analyses reported here provide longitudinal evidence that poorer hearing, greater loneliness, and poorer cognitive performance are meaningfully linked in later life, with loneliness partly accounting for the hearing–cognition association at the Wave-7-aligned level in SHARE. Evidence for mediation in coordinated rates of change was weaker and sensitive to model specification. Hearing-aid exposure was not associated with longer-term loneliness trajectories, but current use was associated with a weaker concurrent association between poorer hearing and loneliness in repeated-measures models. By contrast, genetic analyses did not support large direct effects of hearing phenotypes or loneliness on dementia outcomes with currently available instruments, and instead highlighted neuroticism as an important component of shared genetic architecture. Taken together, these findings suggest that the longitudinal associations may reflect later-life, context-dependent processes that are not easily captured by lifetime genetic liability, but they may also partly reflect residual confounding or broader shared vulnerability. Overall, the results support a meaningful late-life link among hearing difficulty, loneliness, and cognitive performance, while indicating that the underlying causal structure is likely more complex than a single simple mediation pathway.

### Transparency Statement

This study was preregistered on the Open Science Framework (OSF; https://osf.io/xqdhc/overview). We adhered closely to the preregistered analysis plan, and all primary hypotheses, exposures, and outcomes were analyzed as specified. A small number of analytic refinements and clarifications were implemented to address data structure and model convergence issues; these deviations did not change the substantive conclusions and are documented in detail in Supplementary Method 3. The analytic code used in this study will be made available upon publication. Data availability is subject to the access policies of the original data sources, including SHARE and the GWAS datasets used in the genetic analyses.

## Data Availability

The analytic code used in this study will be made available upon publication. Data availability is subject to the access policies of the original data sources, including the Survey of Health, Ageing and Retirement in Europe (SHARE) and the GWAS datasets used in the genetic analyses. SHARE data are available to eligible researchers through the data provider's application procedures, and GWAS summary statistics are available from the original studies, consortia, or repositories subject to their respective access policies.

