## Supplementary Material for "Loneliness as a Pathway Linking Hearing Decline to Cognitive Aging: Longitudinal and Genetic Evidence"

**Supplementary** **Table S1.** **Sample Sizes for Key Variables Across Different Waves in LGCM Models** *(N represents the number of participants available for each variable at each wave.)*

| Variable | Wave 1 | Wave 2 | Wave 3 | Wave 4 | Wave 5 | Wave 6 | Wave 7 | Wave 8 | Wave 9 |
| --- | --- | --- | --- | --- | --- | --- | --- | --- | --- |
| HL | 30,273 | 37,019 | - | 57,745 | 65,869 | 67,940 | 13,939 | - | - |
| Loneliness | - | - | - | - | - | - | 13,566 | 52,393 | 67,706 |
| Objective CO | - | - | - | - | - | - | 74,278 | 52,190 | 67,313 |

Note: Missing values indicate that data for the corresponding variable was not collected or is unavailable for that wave, or is unused in this study.

**Supplementary** **Table S2. Primary univariable two-sample MR (IVW) for loneliness and hearing phenotypes on cognitive and clinical outcomes**

| Comparison (Exposure → Outcome) | nsnp | β (log OR or SD units) | SE | p-value | Bonferroni p (p_bonf) | FDR q (p_fdr) |
| --- | --- | --- | --- | --- | --- | --- |
| Loneliness → Cognitive performance | 14 | 0.0179 | 0.1662 | 0.9141 | 1.0000 | 0.9257 |
| Loneliness → Alzheimer’s disease | 13 | 0.3688 | 0.4394 | 0.4013 | 1.0000 | 0.8026 |
| Loneliness → Dementia (FinnGen R12) | 13 | 0.2374 | 0.2638 | 0.3680 | 1.0000 | 0.8026 |
| SNHL → Cognitive performance | 29 | 0.0189 | 0.0158 | 0.2324 | 1.0000 | 0.8026 |
| SNHL → Alzheimer’s disease | 28 | 0.0050 | 0.0539 | 0.9257 | 1.0000 | 0.9257 |
| SNHL → Dementia (FinnGen R12) | 28 | 0.0386 | 0.0374 | 0.3019 | 1.0000 | 0.8026 |
| HDBM → Cognitive performance | 33 | 0.0865 | 0.1376 | 0.5295 | 1.0000 | 0.8523 |
| HDBM → Alzheimer’s disease | 32 | −0.3812 | 0.4014 | 0.3422 | 1.0000 | 0.8026 |
| HDBM → Dementia (FinnGen R12) | 30 | 0.2711 | 0.4983 | 0.5864 | 1.0000 | 0.8523 |
| ARHI → Cognitive performance | 29 | 0.0226 | 0.0482 | 0.6392 | 1.0000 | 0.8523 |
| ARHI → Alzheimer’s disease | 28 | 0.0470 | 0.1913 | 0.8058 | 1.0000 | 0.9257 |
| ARHI → Dementia (FinnGen R12) | 26 | −0.1834 | 0.2080 | 0.3777 | 1.0000 | 0.8026 |

*Note: All p-values are IVW estimates; Bonferroni correction assumes 12 primary tests.*

**Supplementary Table S3. Two-step mediation MR: loneliness as mediator of hearing loss effects on outcomes (unadjusted for neuroticism)**

| Pathway (Exposure → Loneliness → Outcome) | Proportion mediated | SE | 95% CI lower | 95% CI upper | z | p-value |
| --- | --- | --- | --- | --- | --- | --- |
| SNHL → Loneliness → Cognitive performance | 0.0043 | 0.0438 | −0.0815 | 0.0900 | 0.098 | 0.9222 |
| SNHL → Loneliness → Alzheimer’s disease | 0.3311 | 3.8401 | −7.1953 | 7.8575 | 0.086 | 0.9313 |
| HDBM → Loneliness → Dementia (FinnGen R12) | 0.2889 | 0.6383 | −0.9621 | 1.5399 | 0.453 | 0.6508 |
| ARHI → Loneliness → Dementia (FinnGen R12) | 0.0656 | 0.1190 | −0.1676 | 0.2988 | 0.551 | 0.5816 |

**Supplementary Table S4. Union-of-instruments multivariable MR: SNHL, HDBM, ARHI and loneliness jointly predicting cognitive and clinical outcomes**

| Model ID | Exposure | Outcome (ID) | nsnp | β | SE | p-value | nsnps_model |
| --- | --- | --- | --- | --- | --- | --- | --- |
| SNHL_Union_Lon_to_DemR12 | Loneliness | Dementia_R12 | 15 | −0.1692 | 0.1974 | 0.3912 | 43 |
| SNHL_Union_Lon_to_DemR12 | SNHL | Dementia_R12 | 28 | 0.0521 | 0.0507 | 0.3039 | 43 |
| SNHL_Union_Lon_to_CogPerf | Loneliness | Cognitive performance (ebi-a-GCST006572) | 14 | −0.0167 | 0.1091 | 0.8786 | 39 |
| SNHL_Union_Lon_to_CogPerf | SNHL | Cognitive performance (ebi-a-GCST006572) | 25 | 0.0296 | 0.0283 | 0.2959 | 39 |
| SNHL_Union_Lon_to_AD | Loneliness | Alzheimer’s disease (ieu-b-2) | 13 | −0.2666 | 0.3026 | 0.3782 | 37 |
| SNHL_Union_Lon_to_AD | SNHL | Alzheimer’s disease (ieu-b-2) | 24 | −0.0111 | 0.0826 | 0.8930 | 37 |
| HDBM_Union_Lon_to_DemR12 | HDBM | Dementia_R12 | 31 | 0.3768 | 0.5094 | 0.4594 | 43 |
| HDBM_Union_Lon_to_DemR12 | Loneliness | Dementia_R12 | 14 | 0.0091 | 0.2622 | 0.9722 | 43 |
| HDBM_Union_Lon_to_CogPerf | HDBM | Cognitive performance (ebi-a-GCST006572) | 31 | 0.1535 | 0.2027 | 0.4487 | 43 |
| HDBM_Union_Lon_to_CogPerf | Loneliness | Cognitive performance (ebi-a-GCST006572) | 13 | 0.0540 | 0.1050 | 0.6068 | 43 |
| HDBM_Union_Lon_to_AD | HDBM | Alzheimer’s disease (ieu-b-2) | 30 | −0.4962 | 0.4239 | 0.2418 | 41 |
| HDBM_Union_Lon_to_AD | Loneliness | Alzheimer’s disease (ieu-b-2) | 12 | −0.1029 | 0.2257 | 0.6485 | 41 |
| ARHI_Union_Lon_to_DemR12 | ARHI | Dementia_R12 | 23 | −0.1435 | 0.2360 | 0.5432 | 36 |
| ARHI_Union_Lon_to_DemR12 | Loneliness | Dementia_R12 | 13 | −0.1557 | 0.2405 | 0.5174 | 36 |
| ARHI_Union_Lon_to_CogPerf | ARHI | Cognitive performance (ebi-a-GCST006572) | 26 | 0.0054 | 0.0962 | 0.9549 | 39 |
| ARHI_Union_Lon_to_CogPerf | Loneliness | Cognitive performance (ebi-a-GCST006572) | 13 | −0.0364 | 0.1098 | 0.7403 | 39 |
| ARHI_Union_Lon_to_AD | ARHI | Alzheimer’s disease (ieu-b-2) | 25 | 0.1524 | 0.2542 | 0.5489 | 38 |
| ARHI_Union_Lon_to_AD | Loneliness | Alzheimer’s disease (ieu-b-2) | 13 | −0.3713 | 0.2852 | 0.1929 | 38 |

**Supplementary Table S5. MVMR with loneliness and the large meta-analytic neuroticism jointly predicting outcomes**

| Model ID | Exposure | Outcome (ID) | nsnp | β | SE | p-value | nsnps_model |
| --- | --- | --- | --- | --- | --- | --- | --- |
| Loneliness_Neuro_Union_to_DemR12 | Loneliness | Dementia_R12 | 9 | 0.1113 | 0.1368 | 0.4161 | 134 |
| Loneliness_Neuro_Union_to_DemR12 | Neuroticism | Dementia_R12 | 125 | −0.0154 | 0.0946 | 0.8705 | 134 |
| Loneliness_Neuro_Union_to_CogPerf | Loneliness | Cognitive performance (ebi-a-GCST006572) | 9 | −0.2280 | 0.0641 | 0.00037 | 135 |
| Loneliness_Neuro_Union_to_CogPerf | Neuroticism | Cognitive performance (ebi-a-GCST006572) | 126 | 0.0018 | 0.0441 | 0.9682 | 135 |
| Loneliness_Neuro_Union_to_AD | Loneliness | Alzheimer’s disease (ieu-b-2) | 8 | −0.1337 | 0.1619 | 0.4089 | 123 |
| Loneliness_Neuro_Union_to_AD | Neuroticism | Alzheimer’s disease (ieu-b-2) | 115 | −0.0720 | 0.1126 | 0.5226 | 123 |

**Supplementary Table S6. MVMR with hearing phenotypes and the large meta-analytic neuroticism jointly predicting loneliness**

| Model ID | Exposure | Outcome | nsnp | β | SE | p-value | nsnps_model |
| --- | --- | --- | --- | --- | --- | --- | --- |
| SNHL_Neuro_Union_to_Loneliness | Neuroticism | Loneliness | 119 | −0.0551 | 0.0535 | 0.3032 | 140 |
| SNHL_Neuro_Union_to_Loneliness | SNHL | Loneliness | 21 | 0.0146 | 0.0298 | 0.6240 | 140 |
| HDBM_Neuro_Union_to_Loneliness | HDBM | Loneliness | 21 | 1.0324 | 0.2030 | 3.68×10⁻⁷ | 142 |
| HDBM_Neuro_Union_to_Loneliness | Neuroticism | Loneliness | 123 | −0.0363 | 0.0478 | 0.4479 | 142 |
| ARHI_Neuro_Union_to_Loneliness | ARHI | Loneliness | 17 | −0.4058 | 0.1114 | 0.00027 | 131 |
| ARHI_Neuro_Union_to_Loneliness | Neuroticism | Loneliness | 115 | 0.0013 | 0.0525 | 0.9795 | 131 |

**Supplementary Table S7. Two-step MR mediation: neuroticism-adjusted loneliness as mediator (meta-analytic neuroticism)**

| Pathway (Exposure → Lon(adj) → Outcome) | Proportion mediated | SE | 95% CI lower | 95% CI upper | z | p-value |
| --- | --- | --- | --- | --- | --- | --- |
| SNHL → Lon(adj) → Cognitive performance | −0.1761 | 0.3915 | −0.9435 | 0.5912 | −0.450 | 0.6528 |
| SNHL → Lon(adj) → Alzheimer’s disease | −0.3886 | 4.2671 | −8.7521 | 7.9749 | −0.091 | 0.9274 |
| HDBM → Lon(adj) → Dementia (FinnGen R12) | 0.2618 | 0.4455 | −0.6114 | 1.1350 | 0.588 | 0.5568 |
| ARHI → Lon(adj) → Dementia (FinnGen R12) | 0.2462 | 0.4173 | −0.5717 | 1.0641 | 0.590 | 0.5552 |

**Supplementary Table S8. MVMR with loneliness and GPC neuroticism (sensitivity analyses)**

| Model ID | Exposure | Outcome (ID) | nsnp | β | SE | p-value | nsnps_model |
| --- | --- | --- | --- | --- | --- | --- | --- |
| Loneliness_Neuro_GPC_Union_to_DemR12 | Loneliness | Dementia_R12 | 15 | 0.1366 | 0.3616 | 0.7055 | 16 |
| Loneliness_Neuro_GPC_Union_to_DemR12 | Neuro_GPC | Dementia_R12 | 1 | −0.0956 | 0.6041 | 0.8743 | 16 |
| Loneliness_Neuro_GPC_Union_to_CogPerf | Loneliness | Cognitive performance (ebi-a-GCST006572) | 14 | 0.0352 | 0.2038 | 0.8629 | 15 |
| Loneliness_Neuro_GPC_Union_to_CogPerf | Neuro_GPC | Cognitive performance (ebi-a-GCST006572) | 1 | 0.0534 | 0.3221 | 0.8684 | 15 |
| Loneliness_Neuro_GPC_Union_to_AD | Loneliness | Alzheimer’s disease (ieu-b-2) | 13 | 0.0792 | 0.5179 | 0.8784 | 14 |
| Loneliness_Neuro_GPC_Union_to_AD | Neuro_GPC | Alzheimer’s disease (ieu-b-2) | 1 | −0.8187 | 0.7894 | 0.2997 | 14 |

**Supplementary Table S9. MVMR with hearing phenotypes and GPC neuroticism jointly predicting loneliness**

| Model ID | Exposure | Outcome | nsnp | β | SE | p-value | nsnps_model |
| --- | --- | --- | --- | --- | --- | --- | --- |
| HDBM_Neuro_GPC_Union_to_Loneliness | HDBM | Loneliness | 31 | 0.2912 | 0.1719 | 0.0904 | 32 |
| HDBM_Neuro_GPC_Union_to_Loneliness | Neuro_GPC | Loneliness | 1 | 0.1364 | 0.1197 | 0.2546 | 32 |
| SNHL_Neuro_GPC_Union_to_Loneliness | Neuro_GPC | Loneliness | 1 | 0.0102 | 0.1112 | 0.9272 | 27 |
| SNHL_Neuro_GPC_Union_to_Loneliness | SNHL | Loneliness | 26 | −0.0090 | 0.0172 | 0.6009 | 27 |
| ARHI_Neuro_GPC_Union_to_Loneliness | ARHI | Loneliness | 26 | −0.0426 | 0.0453 | 0.3476 | 27 |
| ARHI_Neuro_GPC_Union_to_Loneliness | Neuro_GPC | Loneliness | 1 | 0.1154 | 0.0689 | 0.0938 | 27 |

**Supplementary Table S10. Two-step MR mediation: neuroticism-adjusted loneliness as mediator (GPC neuroticism; sensitivity analyses)**

| Pathway (Exposure → Lon(adj) → Outcome) | Proportion mediated | SE | 95% CI lower | 95% CI upper | z | p-value |
| --- | --- | --- | --- | --- | --- | --- |
| SNHL → Lon(adj) → Cognitive performance | −0.0167 | 0.1030 | −0.2187 | 0.1852 | −0.162 | 0.8710 |
| SNHL → Lon(adj) → Alzheimer’s disease | −0.1419 | 1.8016 | −3.6730 | 3.3893 | −0.079 | 0.9372 |
| HDBM → Lon(adj) → Dementia (FinnGen R12) | 0.0907 | 0.2673 | −0.4333 | 0.6146 | 0.339 | 0.7346 |
| ARHI → Lon(adj) → Dementia (FinnGen R12) | 0.0317 | 0.0973 | −0.1590 | 0.2224 | 0.326 | 0.7446 |

**Supplementary Table S11. Univariable MR: bidirectional associations between neuroticism, loneliness, and outcomes**

| Pathway (Exposure → Outcome) | Exposure GWAS | Outcome ID | β | SE | p-value | nsnp |
| --- | --- | --- | --- | --- | --- | --- |
| Neuroticism → Loneliness | neuroticism_meta | loneliness | 0.5672 | 0.0236 | 6.63×10⁻¹²⁸ | 118 |
| Loneliness → Neuroticism | loneliness | neuroticism | 0.4455 | 0.0436 | 1.52×10⁻²⁴ | 13 |
| Neuroticism → Dementia (FinnGen R12) | neuroticism_meta | KRA_PSY_DEMENTIA_R12 | 0.0923 | 0.1034 | 0.3719 | 116 |
| Neuroticism → Cognitive performance | neuroticism_meta | ebi-a-GCST006572 | −0.2391 | 0.0395 | 1.41×10⁻⁹ | 128 |
| Neuroticism → Alzheimer’s disease | neuroticism_meta | ieu-b-2 | −0.0694 | 0.1128 | 0.5387 | 117 |

**Supplementary Table S12. Two-step MR mediation with hearing loss as mediator (Loneliness → Hearing loss → Outcomes)**

| Path ID | Exposure (A) | Mediator (M) | Outcome (Y) | Proportion mediated | SE | 95% CI lower | 95% CI upper | z | p-value |
| --- | --- | --- | --- | --- | --- | --- | --- | --- | --- |
| Lon_SNHL_CogPerf | Loneliness | SNHL | Cognitive performance | 0.0499 | 0.4982 | −0.9266 | 1.0264 | 0.100 | 0.9202 |
| Lon_SNHL_AD | Loneliness | SNHL | Alzheimer’s disease | 0.0006 | 0.0073 | −0.0137 | 0.0150 | 0.088 | 0.9299 |
| Lon_SNHL_DemR12 | Loneliness | SNHL | Dementia_R12 | 0.0077 | 0.0299 | −0.0509 | 0.0662 | 0.257 | 0.7974 |
| Lon_HDBM_CogPerf | Loneliness | HDBM | Cognitive performance | 0.6173 | 5.8108 | −10.7716 | 12.0062 | 0.106 | 0.9154 |
| Lon_HDBM_AD | Loneliness | HDBM | Alzheimer’s disease | −0.1321 | 0.2123 | −0.5482 | 0.2840 | −0.622 | 0.5337 |
| Lon_HDBM_DemR12 | Loneliness | HDBM | Dementia_R12 | 0.1460 | 0.3153 | −0.4720 | 0.7639 | 0.463 | 0.6434 |
| Lon_ARHI_CogPerf | Loneliness | ARHI | Cognitive performance | −0.2175 | 2.0708 | −4.2763 | 3.8413 | −0.105 | 0.9164 |
| Lon_ARHI_AD | Loneliness | ARHI | Alzheimer’s disease | −0.0220 | 0.0934 | −0.2050 | 0.1611 | −0.235 | 0.8140 |
| Lon_ARHI_DemR12 | Loneliness | ARHI | Dementia_R12 | 0.1332 | 0.2150 | −0.2882 | 0.5546 | 0.619 | 0.5357 |

**Supplementary Table S13. Genomic regions with strong evidence of colocalization (PP4 ≥ 0.80)**

| Trait 1 | Trait 2 | Chr | Lead position (bp) | PP4 (best prior) | SNPs in region |
| --- | --- | --- | --- | --- | --- |
| HDBM | Cognitive performance | 6 | 30,441,303 | 1.000 | 1,404 |
| Neuroticism | HDBM | 17 | 2,574,821 | 1.000 | 3,288 |
| Neuroticism | ARHI | 17 | 2,574,821 | 0.999 | 3,740 |
| HDBM | Cognitive performance | 6 | 29,346,329 | 0.999 | 1,685 |
| Neuroticism | Cognitive performance | 8 | 143,316,970 | 0.997 | 4,144 |
| Neuroticism | Loneliness | 2 | 226,360,417 | 0.992 | 2,549 |
| HDBM | Loneliness | 18 | 53,252,388 | 0.991 | 1,990 |
| Neuroticism | Loneliness | 9 | 120,496,387 | 0.991 | 2,748 |
| Neuroticism | Cognitive performance | 1 | 75,809,970 | 0.983 | 3,387 |
| Neuroticism | HDBM | 7 | 97,718,650 | 0.980 | 3,255 |
| Neuroticism | Loneliness | 14 | 72,284,548 | 0.976 | 2,573 |
| Neuroticism | Cognitive performance | 5 | 87,978,252 | 0.963 | 2,314 |
| Neuroticism | HDBM | 17 | 44,787,312 | 0.961 | 1,379 |
| Neuroticism | Loneliness | 18 | 50,753,741 | 0.951 | 3,573 |
| Neuroticism | Cognitive performance | 2 | 144,215,811 | 0.943 | 3,040 |
| HDBM | Cognitive performance | 6 | 31,538,497 | 0.943 | 2,447 |
| Loneliness | Cognitive performance | 11 | 47,523,214 | 0.931 | 1,629 |
| Neuroticism | Loneliness | 14 | 69,704,553 | 0.927 | 2,495 |
| HDBM | Cognitive performance | 6 | 26,582,035 | 0.926 | 2,926 |
| Neuroticism | Loneliness | 7 | 12,263,546 | 0.920 | 4,760 |
| Neuroticism | Loneliness | 11 | 13,281,327 | 0.911 | 3,092 |
| Neuroticism | HDBM | 1 | 37,198,130 | 0.910 | 2,758 |
| Neuroticism | Cognitive performance | 16 | 7,664,102 | 0.910 | 7,323 |
| Neuroticism | Cognitive performance | 16 | 87,446,053 | 0.904 | 5,592 |
| Neuroticism | Loneliness | 15 | 78,025,420 | 0.899 | 2,940 |
| Neuroticism | Cognitive performance | 17 | 44,787,312 | 0.899 | 1,355 |
| Neuroticism | HDBM | 6 | 98,521,600 | 0.893 | 2,596 |
| Neuroticism | Loneliness | 12 | 118,796,085 | 0.892 | 2,903 |
| Neuroticism | ARHI | 17 | 44,787,312 | 0.890 | 1,478 |
| Neuroticism | Loneliness | 5 | 120,013,798 | 0.882 | 3,976 |
| Neuroticism | Loneliness | 1 | 75,809,970 | 0.873 | 2,651 |
| Neuroticism | Cognitive performance | 6 | 33,359,820 | 0.870 | 3,626 |
| Neuroticism | Loneliness | 2 | 148,555,489 | 0.868 | 2,055 |
| Neuroticism | Cognitive performance | 14 | 30,268,871 | 0.865 | 3,010 |
| Neuroticism | HDBM | 6 | 27,866,384 | 0.865 | 3,388 |
| HDBM | Cognitive performance | 6 | 27,846,744 | 0.860 | 3,314 |
| Neuroticism | Cognitive performance | 3 | 52,546,820 | 0.856 | 2,696 |
| Neuroticism | Loneliness | 6 | 11,980,752 | 0.856 | 3,211 |
| Neuroticism | Loneliness | 3 | 16,866,253 | 0.855 | 3,070 |
| Neuroticism | Cognitive performance | 5 | 166,179,133 | 0.848 | 3,184 |
| HDBM | Cognitive performance | 17 | 44,221,836 | 0.840 | 586 |
| Loneliness | Cognitive performance | 17 | 44,344,988 | 0.835 | 424 |
| Neuroticism | Cognitive performance | 16 | 5,788,583 | 0.829 | 6,214 |
| Neuroticism | ARHI | 5 | 107,776,094 | 0.827 | 3,850 |
| Neuroticism | Loneliness | 6 | 31,052,632 | 0.824 | 10,599 |
| Neuroticism | Cognitive performance | 6 | 126,792,095 | 0.808 | 1,887 |
| Neuroticism | Loneliness | 16 | 7,664,102 | 0.804 | 5,778 |

*Note*: Posterior probabilities (PP4) are from the best-fitting prior setting (all “Default”; p₁ = p₂ = 1×10⁻⁴, p₁₂ = 1×10⁻⁵).

**Supplementary Methods 1. Longitudinal modeling, hearing-aid moderation, and Mendelian randomization details**

**1. Longitudinal sample construction and missingness**

We constructed a de-duplicated person-by-wave panel from harmonized SHARE data and then created a person-level wide dataset for structural equation modeling. To ensure that growth parameters were estimable for each domain, we required at least two non-missing observations per process: at least two valid observations for hearing across Waves 1, 2, and 4–7 (HL ≥ 2), at least two observations for loneliness across Waves 7–9 (Lon ≥ 2), and at least two observations for objective cognition across Waves 7–9 (ObjCO ≥ 2). Hearing at Wave 3 was set to missing because of documented incompatibilities in wording and response format and was excluded from the hearing growth indicators.

All LGCMs were estimated using full-information maximum likelihood (FIML) under a missing-at-random assumption, allowing individuals with partially observed trajectories to contribute available information. For stability in structural equation modeling with covariates treated as fixed exogenous predictors, we required complete covariate information (age, sex, income, and neuroticism) in the analytic LGCM sample.

**2. Additional longitudinal measurement details**

Observed indicators for hearing, loneliness, and cognition entered the LGCM in their original raw units; indicators were not z-standardized before model estimation.

For the longitudinal covariates, Wave 7 values were prioritized when available to align with the baseline wave for loneliness and cognition. If a Wave 7 covariate value was unavailable, the earliest non-missing value was used as a fallback. Household income was log-transformed as log(1 + income) to reduce skewness. Age, transformed income, and neuroticism were z-standardized prior to modeling.

**3. Time metric and intercept alignment**

To accommodate unequal spacing between SHARE waves, we constructed an empirical time metric using interview timing. For each wave, we computed an average interview date as interview_year + (interview_month / 12); when interview_month was missing, we set it to mid-year for time-score construction. Time scores were defined in years relative to Wave 7. For hearing, time was centered so that Wave 7 = 0 and earlier waves took negative values. For loneliness and cognition, Wave 7 was also set to 0 and subsequent waves reflected elapsed years since Wave 7. Time scores were not min–max scaled; therefore, latent slopes represent change in the original outcome units per year.

To ensure that latent intercepts corresponded to expected levels at the aligned baseline (t = 0, Wave 7), observed indicator intercepts were fixed to zero so that latent intercepts represent the model-implied level at Wave 7 under the centered time metric.

**4. Growth model specification**

We specified a parallel-process latent growth curve model comprising three latent growth processes: hearing, loneliness, and objective cognition. For each domain, we modeled a latent intercept and a latent linear slope, with slope factor loadings fixed to the empirical time scores described above so that slopes represent change per year in the original measurement units. Within each domain, we estimated intercept–slope covariances to allow baseline level and rate of change to covary.

To preserve model parsimony and identifiability, we did not specify cross-domain residual correlations among observed indicators in the primary model; indicator residuals were constrained to be uncorrelated across processes unless explicitly relaxed in sensitivity analyses.

The primary model evaluated mediation at both the aligned level and the change level, while adjusting for covariates. At the aligned level, the loneliness intercept was regressed on the hearing intercept and covariates, and the cognition intercept was regressed on the loneliness intercept, the hearing intercept, and covariates. The aligned-level indirect effect was defined as the product of the hearing-to-loneliness and loneliness-to-cognition coefficients, and the aligned-level total effect was defined as the sum of the direct hearing-to-cognition effect and the indirect effect.

At the change level, the loneliness slope was regressed on the hearing slope and covariates, and the cognition slope was regressed on the loneliness slope, the hearing slope, and covariates. The slope indirect effect was defined as the product of the hearing-slope-to-loneliness-slope and loneliness-slope-to-cognition-slope coefficients, and the slope total effect was defined as the sum of the direct hearing-slope-to-cognition-slope effect and the indirect effect.

Covariates were included as predictors of both the aligned-level and slope-level structural equations for loneliness and cognition.

**5. Sensitivity models**

To assess robustness of slope-mediation inferences to alternative handling of baseline levels, we fit two prespecified sensitivity models.

**Sensitivity Model A (slope-only with correlated intercepts).**
We retained the slope mediation structure while allowing intercepts for hearing, loneliness, and cognition to correlate and removing intercept regressions across domains.

**Sensitivity Model B (level-only with correlated slopes).**
We retained the aligned-level mediation structure while allowing slopes for hearing, loneliness, and cognition to correlate and removing slope regressions across domains.

Sensitivity models that failed to converge were not interpreted. To prevent occasional improper solutions driven by near-zero residual variances in specific observed indicators, we applied small lower-bound constraints to selected indicator residual variances as a numerical safeguard.

**6. Estimation and model fit**

LGCMs were estimated in R (version 4.5.2) using the lavaan package. Models were fitted using maximum likelihood with robust corrections (MLR) and full-information maximum likelihood for missing data under a missing-at-random assumption (missing = "fiml"). The mean structure was included, and models were optimized using a robust optimizer with parameter bounds to improve numerical stability. Model fit was evaluated using the comparative fit index (CFI), Tucker–Lewis index (TLI), root mean square error of approximation (RMSEA), and standardized root mean square residual (SRMR), using robust or scaled versions when available.

**7. Hearing-aid moderation analyses**

Because hearing aids are a clinically relevant management strategy for hearing loss, we conducted four complementary analyses to test whether hearing-aid use moderated the hearing–loneliness association.

Hearing-aid exposure was operationalized in two ways: cumulative hearing-aid use through Wave 7 (HA_cum) and current hearing-aid use at a given wave (HA_it). HA_cum was derived from a cumulative proportion measure across Waves 1–7, prioritizing the Wave 7 value when available; otherwise, we used the last observed value prior to Wave 7, with a fallback to the within-person median across pre-7 waves. Current hearing-aid use was treated as a binary indicator (0/1). Unless otherwise noted, moderation analyses adjusted for age, sex, income, and neuroticism.

**Analysis A.**
We extracted individual factor scores for latent slopes from the fitted primary LGCM, merged these scores with HA_cum and covariates, and estimated a linear regression model in which the loneliness slope was regressed on the hearing slope, HA_cum, and their interaction. The hearing slope and HA_cum were mean-centered prior to forming the interaction term.

**Analysis B.**
We used person-wave data for Waves 7–9 with observed loneliness and constructed time in years since each individual’s Wave 7 interview when interview dates were available, otherwise using wave index as a fallback. We centered time at the sample mean, standardized loneliness, and fit linear mixed-effects models with random intercepts and, when supported, random slopes for time. The fixed-effects portion included time, HA_cum, and their interaction, with covariate adjustment. If the random-slope model was singular, we refit a random-intercept-only model.

**Analysis C.**
We focused on the Wave 7 cross-sectional association and tested whether current hearing-aid use modified the hearing–loneliness association at baseline. Hearing and loneliness at Wave 7 were standardized, and loneliness was regressed on hearing, current hearing-aid use, and their interaction, with covariate adjustment.

**Analysis D.**
We extended the moderation test to all available person-wave observations with non-missing hearing, hearing-aid use, and loneliness by fitting linear mixed-effects models with a random intercept for participant. The fixed-effects portion included standardized hearing, current hearing-aid use, and their interaction. Baseline covariates for Analysis D were defined using each participant’s earliest non-missing value across waves. As robustness checks, we additionally included wave fixed effects and decomposed hearing into between-person and within-person components to test moderation for each component separately. All mixed-effects models were fitted in R using the lme4 package with REML = FALSE.

**8. MR data sources**

We used summary statistics from large-scale GWAS for all exposure and outcome traits. Detailed characteristics of each dataset, including sample sizes, phenotype definitions, and data sources, are provided in Table 1. Briefly, the primary hearing-loss exposure was SNHL diagnosis from FinnGen R12, supplemented by HDBM and ARHI from UKB. Loneliness was instrumented using the largest available meta-analysis (Abdellaoui et al., 2019a). Cognitive outcomes included cognitive performance (COGENT + UKB), AD dementia (IGAP), and all-cause dementia (FinnGen R12). Neuroticism instruments were derived from a recent large GWAS meta-analysis (Gupta et al., 2024) for the primary MVMR analyses. As a sensitivity analysis to reduce possible sample-overlap concerns, key models were repeated using an independent neuroticism GWAS from the GPC.

**9. Genetic instrument selection**

For all exposure traits, we selected SNPs associated with the phenotype at a genome-wide significance threshold of P < 5 × 10⁻⁸. To ensure independence, we clumped variants using an LD threshold of r² < 0.001 within a 10,000 kb window based on the 1000 Genomes Project European reference panel. Effect sizes from outcome GWAS were harmonized to ensure effect alleles were aligned to the same strand. Palindromic SNPs with intermediate allele frequencies or incompatible alleles were excluded to prevent strand ambiguity.

We calculated F-statistics as a diagnostic of instrument strength and weak-instrument risk. We report these diagnostics in the supplementary tables rather than using them as the sole instrument-selection criterion.

**10. Primary MR analyses**

We first conducted primary two-sample MR analyses to estimate total effects between each of the four exposures (SNHL, HDBM, ARHI, and loneliness) and each of the three outcomes (cognitive performance, AD dementia, and all-cause dementia), yielding 12 primary pathways. The IVW method was used as the primary estimator. To account for multiple testing across the 12 primary hypotheses, we applied a Bonferroni correction with a significance threshold of P < 0.004. Results with P < 0.05 but above this threshold were considered nominally significant. FDR correction was also applied for supplementary comparison.

**11. Two-step MR mediation analysis**

To investigate whether loneliness mediated the pathway from hearing phenotypes to cognitive outcomes, we employed a two-step MR framework. We estimated the indirect effect by multiplying the exposure-to-mediator effect by the mediator-to-outcome effect. Standard errors and 95% confidence intervals for the indirect effects were derived using the delta method. To minimize bias from sample overlap, we prioritized three mediation pathways: (1) SNHL → loneliness → cognitive performance, (2) SNHL → loneliness → AD dementia, and (3) HDBM or ARHI → loneliness → all-cause dementia.

**12. MVMR and neuroticism adjustment**

We conducted MVMR to disentangle direct versus shared effects and to account for potential horizontal pleiotropy introduced by correlated traits. We first modeled hearing phenotypes and loneliness jointly against cognitive outcomes and then extended those models to include neuroticism where loneliness was an exposure or mediator.

To ensure robust estimation, we evaluated three alternative strategies for constructing instruments. Our final primary specification used a union-of-instruments approach. For each model, genome-wide significant, LD-clumped SNPs from all relevant exposures were pooled and re-clumped to ensure mutual independence. Association estimates for all exposures and outcomes were then extracted for this union set, allowing all modeled traits to be instrumented symmetrically and maximizing instrument coverage.

**13. Additional MR sensitivity analyses**

We conducted complementary exploratory analyses of bidirectional relationships between neuroticism and loneliness and analyses of neuroticism in relation to the three cognitive outcomes. As a negative-control-style sensitivity analysis, we also explored the reverse mediation pathway (loneliness → hearing phenotype → cognitive outcome).

To assess robustness of IVW findings, we applied the weighted median estimator, the MR-Egger intercept test for directional pleiotropy, Cochran’s Q statistic for heterogeneity, and MR-PRESSO to identify and correct for pleiotropic outliers.

**Supplementary Methods 2. Colocalization implementation details**

This section provides implementation details for the colocalization strategy described in the main Methods, including data parsing, harmonization, locus definition, SNP alignment, prior-sensitivity analyses, and output generation.

All analyses were implemented in R (version 4.5.2) using the coloc package (coloc.abf), with data handling performed using data.table and dplyr. We conducted colocalization across a prespecified set of 22 trait pairs covering the core pathways assessed elsewhere in the manuscript. These included each hearing-loss (HL) phenotype with loneliness, loneliness with each cognitive outcome, each HL phenotype with each cognitive outcome, and neuroticism with each of the other traits (loneliness, three HL phenotypes, and three cognitive outcomes). Traits were grouped into quantitative phenotypes (loneliness, neuroticism, and cognitive performance) and case-control phenotypes (SNHL, HDBM, ARHI, AD dementia, and all-cause dementia). For each trait, we used full GWAS summary statistics rather than restricting analyses to the genetic instruments used in MR.

Sample size parameters were supplied as recorded in the preregistered analytic registry and analysis scripts: SNHL (N = 482,076; case fraction *s* = 44,745/482,076), HDBM (N = 453,482; *s* = 171,586/453,482), ARHI (N = 330,759; *s* = 125,688/330,759), loneliness (N = 511,280), cognitive performance (N = 257,841), AD dementia (N = 63,926; *s* = 21,982/63,926), all-cause dementia (N = 383,461; *s* = 21,157/383,461), and neuroticism (N = 682,688).

**1. Summary-statistics parsing and harmonization**

Summary statistics were ingested from either plain-text GWAS files or OpenGWAS-style VCFs and then standardized to a common schema containing chromosome (chr), base-pair position (pos), SNP identifier (snp), effect size (beta), variance of the effect estimate (varbeta = SE²), p-value (p), and minor allele frequency (MAF, when available).

For text-based inputs, we harmonized heterogeneous column labels by mapping common variants of SNP ID (e.g., rsid, rsids, variant), effect size (beta, b, logOR), standard error (se), p-value (p, pval, p_value), chromosome, position, and allele frequency to the unified schema. When a formal rsID column was unavailable, we constructed a positional identifier using the concatenation CHR:POS to ensure the presence of a unique key for downstream matching.

For VCF-based inputs, we parsed the CHROM, POS, and ID fields into chr, pos, and snp, and extracted association statistics from the VCF FORMAT/STAT field as encoded in the OpenGWAS export. Effect estimates and standard errors were read as numeric fields, and p-values were recovered from the stored −log10(p) statistic using p = 10^(−x). Minor allele frequency was retained when available. Across all sources, variants with missing beta, standard error, or p-value were removed, and varbeta was computed as SE².

**2. Within-trait deduplication**

To preserve one-variant-one-row matching within each trait before cross-trait harmonization, we enforced a one-row-per-variant structure for every trait-specific GWAS dataset. When duplicate variant identifiers occurred within a trait, for example because of repeated rsIDs, repeated CHR:POS entries arising from multi-allelic sites, imputation artifacts, or concatenation across partitions, we retained a single record per variant.

Variants were keyed by SNP ID when available and otherwise by genomic position (CHR:POS). When duplicates were present, we resolved them deterministically by retaining the row with the smallest p-value. If p-values were identical or unavailable, we retained the row with non-missing beta and standard error; otherwise, we retained the first occurrence after sorting. This step ensured that subsequent locus extraction and SNP alignment did not produce ambiguous matches or inflate overlap counts.

**3. Locus definition and regional extraction**

Because coloc.abf is defined for a genomic region, we implemented a two-stage regional testing procedure.

First, for each trait used to seed loci in a given trait pair, we identified genome-wide significant variants using p < 5 × 10⁻⁸. We then selected approximately independent lead variants via a greedy distance-based clumping algorithm in physical space. Specifically, significant variants were ordered by ascending p-value, the top variant was retained as a lead SNP, and all remaining significant variants within ±1 Mb on the same chromosome were removed. This process was repeated iteratively until no significant variants remained. If the first trait in a pair contained no genome-wide significant variants, we repeated the same procedure using the second trait to obtain lead loci. Each retained lead SNP defined the center of a candidate region for colocalization testing.

Second, for each lead SNP, we extracted a ±500 kb window around the lead position from both trait datasets using chromosome and base-pair position. Within each region, we restricted analysis to variants present in both datasets by taking the intersection of SNP identifiers. We required at least 50 overlapping variants within the window to proceed, because sparse overlap can destabilize posterior probability estimation and make discrimination between PP₃ and PP₄ unreliable. When the overlap criterion was not met, the region was excluded from testing and no colocalization posterior probabilities were generated for that locus. This overlap-based gatekeeping also served as a conservative quality-control mechanism when trait pairs had limited cross-dataset compatibility, for example because of differences in genomic build or variant representation.

**4. Regional SNP alignment and coloc inputs**

For each retained region, we constructed separate coloc input objects for the two traits. For each trait, we supplied vectors of beta and varbeta named by SNP ID, the SNP identifier list, and the corresponding base-pair positions. Quantitative traits were specified with type = "quant" and total sample size N. Case-control traits were specified with type = "cc", total sample size N, and case fraction s. When MAF was available and non-missing for a region, it was included in the coloc input to improve calibration.

Before calling coloc.abf, we enforced exact SNP alignment by intersecting on SNP IDs, removing any remaining duplicates, sorting both regional datasets by SNP ID, and verifying that SNP ordering was identical across the two traits. Analyses were run only after this one-to-one, order-consistent alignment check was satisfied.

**5. Prior-sensitivity analyses**

To assess robustness to prior assumptions, we performed a prior-sensitivity analysis for every tested region by repeating coloc.abf under two prior settings. In the canonical setting, we used p1 = 1 × 10⁻⁴ and p2 = 1 × 10⁻⁴ for the probability that each trait has a causal variant in the region and p12 = 1 × 10⁻⁵ for the probability that both traits share a single causal variant. In a more conservative setting, we held p1 and p2 constant and reduced the shared-variant prior to p12 = 1 × 10⁻⁶, which down-weights colocalization a priori and provides a stricter test for a shared regional signal.

For each region and prior setting, we extracted posterior probabilities for hypotheses H₀ through H₄, along with the number of overlapping SNPs included in the computation.

**6. Output generation and interpretation**

Outputs were written at the trait-pair level and retained at the region-by-prior granularity to support full traceability. For each trait pair, we saved a results table containing the trait identifiers, chromosome, lead position, the number of overlapping SNPs in the region, the prior setting, and posterior probabilities PP₀ through PP₄ from coloc.abf.

These regional outputs were then aggregated for reporting. High-confidence colocalized loci were defined as regions with PP₄ ≥ 0.80 under the default prior setting (p₁ = p₂ = 1 × 10⁻⁴; p₁₂ = 1 × 10⁻⁵), whereas regions with PP₃ > 0.50 and low PP₄ were interpreted as consistent with distinct causal variants within the same locus. This region-level aggregation was used to summarize the colocalization landscape across the prespecified 22 trait pairs and to facilitate comparison with the MR/MVMR and LGCM findings presented in the Results and Discussion sections.

**Supplementary Method 3. Deviations from the Preregistered Analysis Plan**

This project was preregistered on the Open Science Framework (OSF; <https://osf.io/xqdhc/overview>). Below we describe all deviations from the preregistered plan and the rationale for each change.

**1. Exclusion of subjective cognition from the LGCM mediation model**
In the preregistration, we planned to model both objective cognition and subjective cognition as parallel cognitive outcomes in the latent growth curve mediation (LGCM) framework. In practice, the subjective cognition measure was available for only two waves with sufficient data quality, which is insufficient for estimating a stable latent slope factor in LGCM. Attempts to fit a three-process model including subjective cognition (hearing, loneliness, subjective cognition) led to non-identification and unstable parameter estimates. To preserve model identifiability and interpretability, we therefore restricted the LGCM mediation analyses to objective cognition (immediate + delayed word recall across Waves 7–9), as reported in the main text.

**2. Simplification of the LGCM covariate structure**
The preregistered plan specified that we would adjust the LGCM for a broad set of baseline sociodemographic covariates, including education. When we implemented this specification, however, the models showed poor global fit, frequent convergence problems, and indications of over-parameterization (e.g., inadmissible solutions and non–positive definite covariance matrices). To obtain a stable and interpretable model of longitudinal change, we adopted a more parsimonious covariate set consisting of age, sex, income, and neuroticism—variables that were already prioritized in the preregistration as key confounders and stratifiers of interest. All primary LGCM mediation results reported in the manuscript are based on this reduced, well-behaved model.

**3. Addition of MR analyses involving neuroticism**
Our preregistration focused on the causal pathways linking hearing loss, loneliness, and cognitive outcomes. During the analysis, and motivated by prior evidence for a strong genetic and phenotypic overlap between loneliness and neuroticism, we added a set of clearly labeled exploratory MR analyses. Specifically, we examined (a) bidirectional causal effects between neuroticism and loneliness, and (b) the effects of neuroticism on the three cognitive outcomes (cognitive performance, Alzheimer’s disease, and all-cause dementia). These analyses were not part of the preregistered primary hypotheses and are presented as hypothesis-generating results in the Supplementary Material to clarify the specificity of loneliness-related findings.

None of these deviations alter the core design of the study. The primary LGCM mediation analyses of hearing loss → loneliness → objective cognition, the main two-sample MR pathways between hearing phenotypes, loneliness, and cognitive outcomes, and the overarching interpretation of results remain consistent with the preregistered plan.
